# Trust as a Hidden Driver of Epidemic Dynamics: A Missing Parameter in Compartmental Disease Transmission Models

**DOI:** 10.64898/2026.06.15.26355705

**Authors:** Alexander Zapf, George Dewey, Katherine Ognyanova, Matthew Baum, William P. Hanage, Marc Lipsitch, Ata A. Uslu, James N. Druckman, Roy Perlis, David Lazer, Mauricio Santillana

## Abstract

Compartmental models of infectious disease transmission make assumptions about human behaviors. Specifically, they parameterize interactions across population groups, assumed to have distinct epidemiologically-relevant behavioral patterns, primarily through contact matrices stratified by demographic variables such as age, gender, or socioeconomic status. Although such demographic characteristics are readily measurable, they may inadequately capture the social and psychological forces that govern protective behaviors. Drawing on 20 waves of a national survey conducted throughout the COVID-19 pandemic in the United States, we show that institutional trust — particularly trust in public health agencies, physicians, and hospitals — is a dominant predictor of protective behavior adoption. For mask wearing during periods of strongest pandemic activity, for example, institutional trust explains more behavioral variance across population groups than age, income, education, and partisan affiliation combined. In unadjusted analyses, the difference in protective behavior adoption between individuals with the highest and lowest trust in the CDC was four- to six-fold larger than the corresponding differences by age, income, or educational attainment, and exceeded the difference between Democratic and Republican respondents. This association was institutionally specific (e.g., the relationship attenuates for trust in banks), and behaviorally specific (e.g., trust in the CDC is associated with protective behaviors but not visiting a doctor). The latter suggests that trust modifies voluntary compliance with public health recommendations rather than access to or use of healthcare. We conclude that compartmental models of disease transmission would be substantially improved by incorporating institutional trust as a stratifying variable. We additionally offer a trust-integrated mathematical modeling framework and recommendations for the data infrastructure needed for its implementation.

## Introduction

The transmission of infectious diseases is fundamentally governed by human behavior. As the world experienced during the COVID-19 pandemic, the pace and eventual magnitude of an epidemic^1,2^ is determined by individuals’ choices to engage in protective behavior, like avoiding contacts, wearing masks, seeking medical care, or accepting vaccination. These individual-level decisions aggregate into population-level contact patterns that modulate the effective reproduction number of a pathogen. By extension, decisions drive the dynamics of epidemic proliferation. Substantial efforts have been invested in incorporating behavioral dynamics into susceptible-infectious-recovered (SIR) compartmental models which simulate the spread of infectious disease through a population^3^. This has primarily involved demographically stratified contact matrices derived from social contact surveys^4–7^, agent-based models, and extended frameworks that couple epidemiological dynamics with behavioral or economic choices. Taken together, these models share a reliance on using measurable demographic variables (e.g., age, gender, or socioeconomic status) to explain variations in epidemic trajectories across population groups.

These approaches face two fundamental limitations. First, they rely on demographic and structural proxies for behavioral heterogeneity rather than direct measures of the social and psychological forces that determine population-level behavioral change during an epidemic. Second, the measures used to determine behavioral heterogeneity are generally static and do not change with respect to the epidemic. During the COVID-19 pandemic, behavioral responses to identical epidemiological conditions diverged sharply across communities, states, and demographic groups in ways that stable (static) characteristics such as age, gender, geography, and income cannot fully explain^8–10^. These divergences tracked consistently with the degree to which individuals trusted physicians, hospitals, and public health agencies such as the US Centers for Disease Control and Prevention (CDC). These institutions form the backbone of response to epidemics in the United States, with physicians and hospitals serving as frontline agents for prevention. Simultaneously, public health agencies and government bodies define policies to reduce the burden of such epidemics on the population-at-large. By trusting these institutions, individuals accept their risk assessments and follow recommendations to adopt costly protective behaviors. Unlike demographic variables—distal to decision-making and primarily static during an epidemic—trust is a proximate factor that evolves with the epidemic, reflecting the bidirectional feedback between transmission and behavior.

Additionally, while partisanship was a significant factor in driving differences in pandemic outcomes across the United States^11^, partisanship is a more static characteristic that does not reflect the rapidly changing nature of many aspects of institutional trust. While existing studies have explored the statistical associations between trust in health institutions and either protective behavior adoption and vaccine uptake^12,13^, these studies only consider demographic variables as controlling factors in regression models and do not consider trust, demographics, protective behavior adoption, or vaccine uptake in the context of epidemiological modeling.

Institutional trust, the confidence individuals place in governmental and public health organizations to act in the public interest, provides a theoretically grounded mechanism linking social context to epidemic-relevant behavior. When individuals trust health institutions, they are more likely to accept their risk assessments as credible and to adopt the protective behaviors those institutions recommend. Consistent with the Health Belief Model^14,15^ trust shapes perceived susceptibility and severity, which in turn drive discretionary compliance: high-trust individuals treat public health guidance as a meaningful signal of personal risk, whereas low-trust individuals are more likely to discount that guidance and engage in risk-exposing behaviors. Indeed, a series of papers highlight trust as a key lever of behaviors during public health crises such as COVID-19^16–20^. We build on this work by measuring institutional trust during the COVID-19 pandemic to complement existing work on trust in non-pandemic scenarios^21–23^ and by highlighting trust as a key driver of behavior beyond partisanship or demographics alone.

Furthermore, despite its demonstrated behavioral relevance, institutional trust has not been incorporated into mathematical disease models as a structural parameter. We propose that including institutional trust in epidemiological models will improve characterization of effective contact rates, clarify subpopulations that drive transmission, and reduce the bias in model predictions. Here, we use data from 20 waves of the Civic Health and Institutions Project (CHIP50)^24^, a nationally representative nonprobability survey, to provide the first systematic empirical characterization of trust during the COVID-19 pandemic, using rigorously adjusted comparisons to test whether institutional trust independently drives protective behavior. We identify larger differences in protective behavior adoption when comparing trust groups to groups stratified by demographics or partisanship which hold even after adjustment for demographic, political, and geographic confounders. We also emphasize the robustness of our conclusions by using several control methods to argue that the association between institutional trust and behavior adoption reflects institution-specific trust rather than a generic disposition or a proxy for underlying structural disparities. Finally, we show the importance of understanding institutional trust alongside conventional sociodemographic predictors and propose a mathematical framework for incorporating trust into behavior-coupled compartmental models.

## Results

### Trust consistently produces larger differences in population-level protective behavior adoption than demographics

Across all 20 survey waves and all four protective behaviors — mask wearing, avoiding contact with others, avoiding crowded places, and frequent handwashing — trust in the CDC was associated with the largest and most temporally consistent between-group adherence differences of any stratifier examined (age, gender, race/ethnicity, income, education, ideology and political leaning; **Figures 1–2**, **Supplementary Figures S1 – S8**). For conciseness, we focus on trust in the CDC in the main text; analogous analyses for trust in other health institutions, including doctors and hospitals (sources of routine care) and state governments (i.e., institutions which direct public health policy), yielded similar behavioral differences and are reported in the Supplementary Materials. The unadjusted trust gaps were substantial and significant for every behavior: the mean difference in adherence between highest-trust (“a lot”) and lowest-trust (“no trust”) respondents was 38.8 pp for masking (**Figure 1**; p < 0.001 in every wave) and ranged from roughly 24 to 39 pp across the four behaviors.

**Fig 1.**
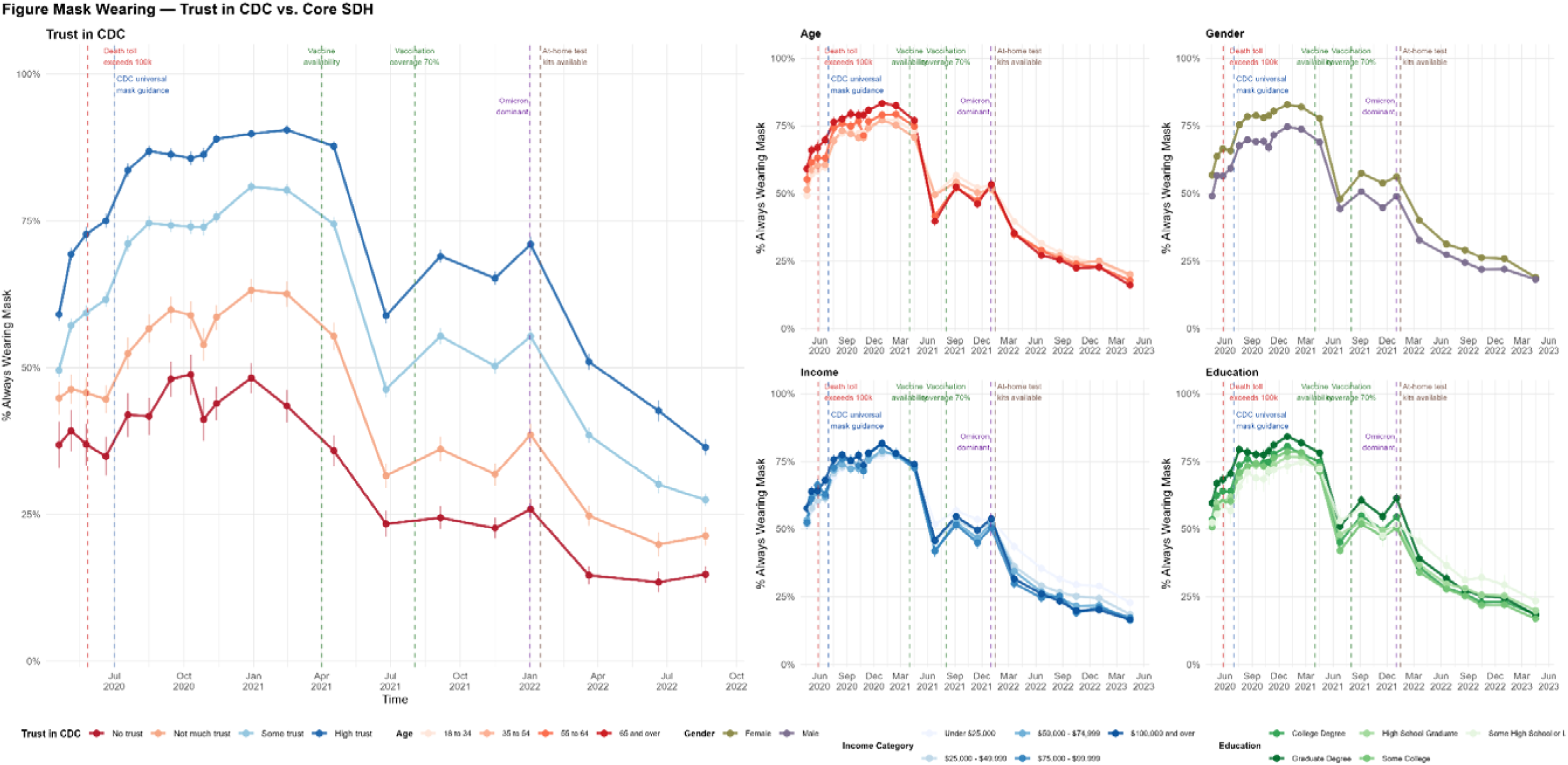
Temporal trajectory of mask wearing, stratified by trust in the CDC and by core sociodemographic predictors (Age, Gender, Income, Education). The left panel shows the temporal evolution of the proportion of survey respondents reporting always wearing a mask in public, stratified by self-reported trust in the CDC (high, some, not much, and none — corresponding to the survey response options “a lot”, “some”, “not too much”, and “not at all”). The four right-hand panels show the same outcome stratified by age, gender, household income, and educational attainment. Points represent survey-weighted estimates and vertical bars denote 95% confidence intervals, based on *20* waves of the CHIP50 Project survey. Dashed vertical lines mark key pandemic milestones with potential impact on protective behaviors. *Death toll exceeds 100k* represents the date the officially recorded COVID-19 death toll in the US exceeded 100,000 deaths (May 27, 2020). *CDC universal mask guidance* (July 2020) marks the US CDC’s recommendation that all individuals use face masks in indoor public settings. *Vaccine availability* (spring 2021) denotes the point at which COVID-19 vaccines became widely available to the US population without the eligibility restrictions imposed by the earlier prioritization strategies, reached for all adults in approximately April 2021. *Vaccination coverage 70%* (summer 2021) marks the point at which at least 70% of US adults had received one or more doses of a COVID-19 vaccine, reached in early August 2021. *Omicron dominant* (January 2022) marks the appearance and rapid rise to dominance of the Omicron (B.1.1.529) variant among US infections. *At-home tests widely available* (January 2022) marks the point at which rapid antigen self-tests became broadly accessible to the US public, coinciding with the launch of the federal free-distribution program and the requirement that insurers cover at-home tests.

**Fig 2.**
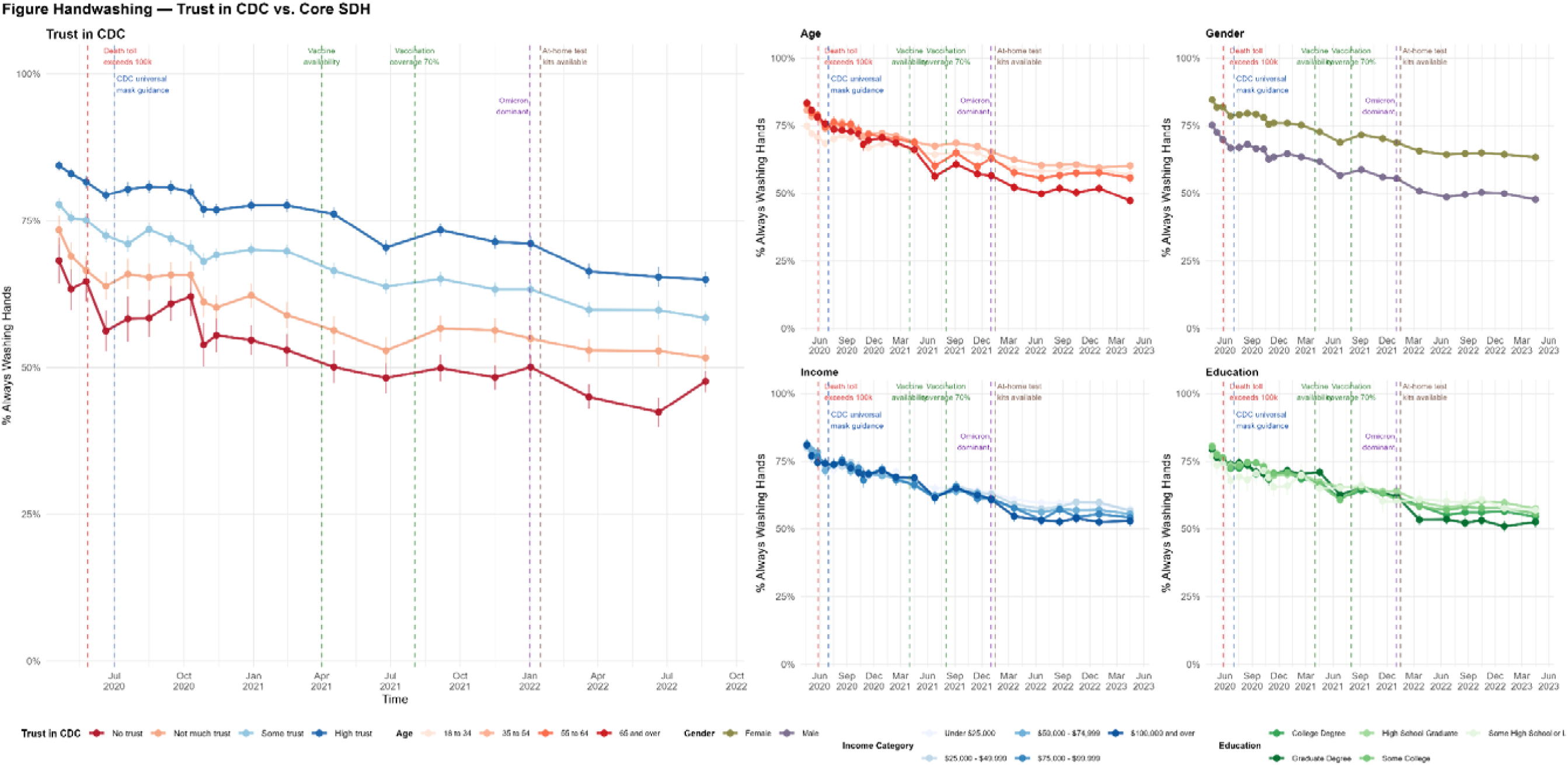
Temporal trajectory of handwashing, stratified by trust in the CDC and by core sociodemographic predictors (Age, Gender, Income, Education). The left panel shows the temporal evolution of the proportion of survey respondents reporting always washing their hands, stratified by self-reported trust in the CDC (high, some, not much, and none — corresponding to the survey response options “a lot”, “some”, “not too much”, and “not at all”). The four right-hand panels show the same outcome stratified by age, gender, household income, and educational attainment. Points represent survey-weighted estimates and vertical bars denote 95% confidence intervals, based on *20* waves of the CHIP50 Project survey. Dashed vertical lines mark key pandemic milestones with potential impact on protective behaviors. *Death toll exceeds 100k* represents the date the officially recorded COVID-19 death toll in the US exceeded 100,000 deaths (May 27, 2020). *CDC universal mask guidance* (July 2020) marks the US CDC’s recommendation that all individuals use face masks in indoor public settings. *Vaccine availability* (spring 2021) denotes the point at which COVID-19 vaccines became widely available to the US population without the eligibility restrictions imposed by the earlier prioritization strategies, reached for all adults in approximately April 2021. *Vaccination coverage 70%* (summer 2021) marks the point at which at least 70% of US adults had received one or more doses of a COVID-19 vaccine, reached in early August 2021. *Omicron dominant* (January 2022) marks the appearance and rapid rise to dominance of the Omicron (B.1.1.529) variant among US infections. *At-home tests widely available* (January 2022) marks the point at which rapid antigen self-tests became broadly accessible to the US public, coinciding with the launch of the federal free-distribution program and the requirement that insurers cover at-home tests.

These trust-related differences consistently dwarfed those produced by conventional demographic stratifiers, which were in the single digits (on average 6–10 pp for age, educational attainment, and household income), and thus four- to six-fold smaller. The trust contrast also exceeded the partisan difference – widely regarded as a dominant driver of COVID-19 behavioral variation^11^ (mean difference was 25.9 pp for masking, Democratic vs. Republican) – in every wave analyzed. This pattern replicated across all measured protective behaviors, including distancing, avoiding crowds, and strict handwashing, as well as for vaccine uptake (**Supplementary Figures S9 – S10**), indicating that trust influences protective behavior more strongly than political identity. However, these disparities reflect unadjusted estimates, so the magnitude of the difference by trust could reflect its correlation with partisanship, socioeconomic position, or other demographic characteristics. These striking raw behavioral differences were therefore further assessed using rigorous, survey-weighted multivariable analyses to differentiate whether institutional trust is an independent driver of protective-behavior adoption or merely a proxy for underlying structural disparities.

### Trust in the CDC remains the dominant behavioral predictor after multivariable adjustment

To assess whether the unadjusted trust differences reported above reflect institutional trust rather than its correlation with sociodemographic and political covariates, we estimated survey-weighted multivariable logistic regressions for each protective behavior within each CHIP50 wave, jointly adjusting for age, race/ethnicity, education, household income, urbanicity, partisan leaning, ideology, U.S. Census region, and employment. For mask wearing and trust in the CDC, the adjusted high- versus no-trust risk differences remained substantial throughout the pandemic, with estimates of 31.7 pp (95% CI 27.9–35.5) in June 2020 (Wave 5, pre-vaccine), 37.2 pp (95% CI 34.2–40.2) in February 2021 (Wave 16, vaccine rollout), and 32.5 pp (30.1–35.0) in January 2022 (Wave 21, Omicron; **Figure 5**). Analogous adjusted differences were observed for always avoiding contact, always avoiding crowds, and frequent handwashing (pooled estimates spanning 13–27 pp; **Figure 5**). In sensitivity analyses removing partisan leaning and ideology from the covariate set, the adjusted mask-wearing contrast increased by 4.6–8.2 pp— to 36.3 pp at Wave 5, 44.2 pp at Wave 16, and 40.7 pp at Wave 21 — indicating that partisanship and ideology absorb only a modest portion of the trust-related variance and that the trust difference remains large and significant in the fully adjusted models (**Figure 5**, cf. **Supplementary Figures S11 – S16**). In a separate set of sensitivity analyses that additionally conditioned on an index of general trust in others, the CDC mask-wearing difference was essentially unchanged and remained significant (31.2 pp [95% CI 26.7–35.7] at Wave 5, 37.0 pp [33.4–40.6] at Wave 16, and 31.2 pp [28.1–34.4] at Wave 21; all p < 0.0001, **Supplementary Figures S17 – S18**). The similarity of these estimates to the primary models indicates that the CDC difference reflects trust specifically in public-health institutions beyond a dispositional propensity toward interpersonal trust.

**Fig 3.**
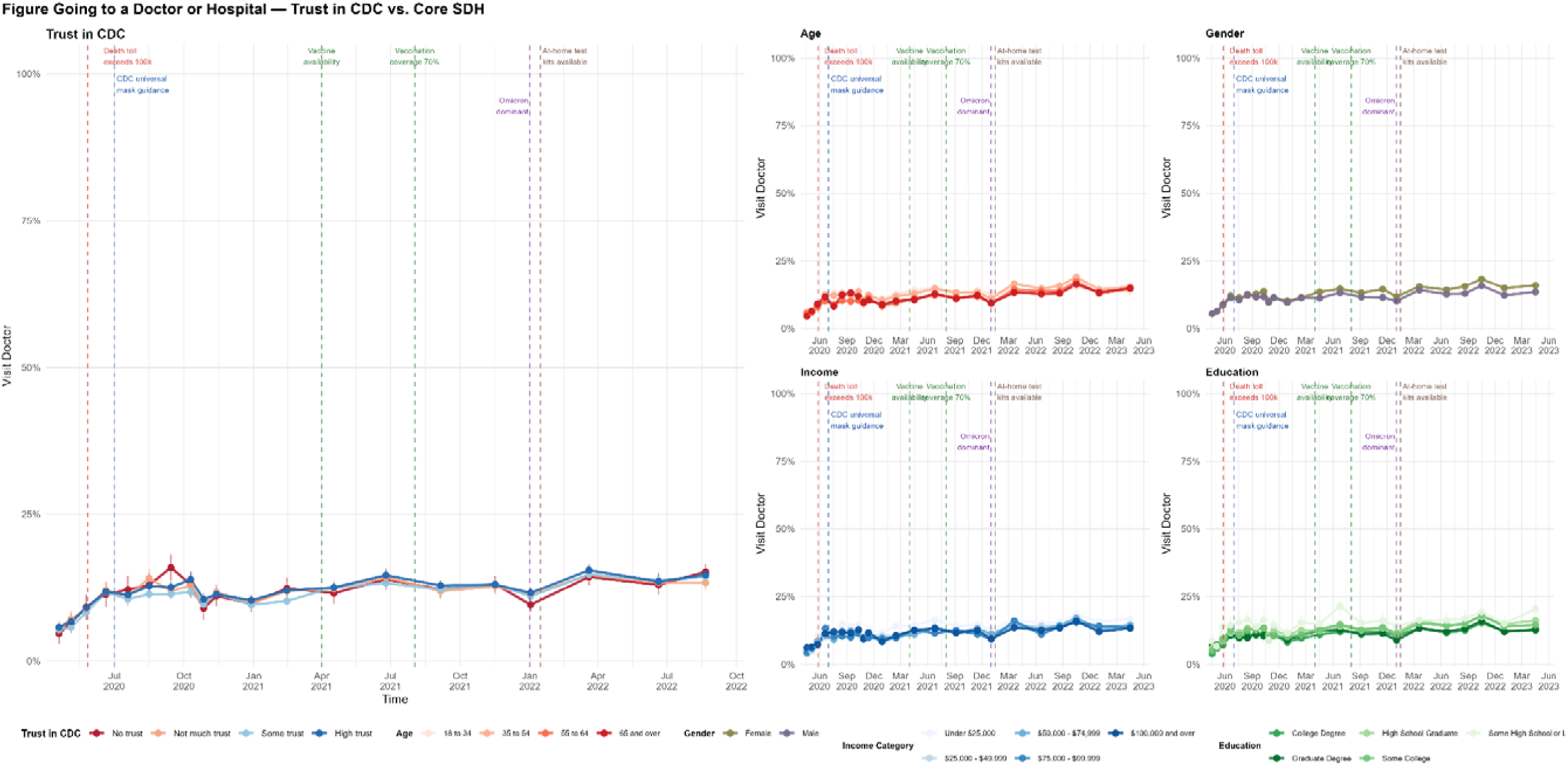
Temporal trajectory of going to a doctor or hospital, stratified by trust in the CDC and by core sociodemographic predictors (Age, Gender, Income, Education). The left panel shows the temporal evolution of the proportion of survey respondents reporting to have visited a doctor or hospital in the last 24 hours, stratified by self-reported trust in the CDC (high, some, not much, and none — corresponding to the survey response options “a lot”, “some”, “not too much”, and “not at all”). The four right-hand panels show the same outcome stratified by age, gender, household income, and educational attainment. Points represent survey-weighted estimates and vertical bars denote 95% confidence intervals, based on *20* waves of the CHIP50 Project survey. Dashed vertical lines mark key pandemic milestones with potential impact on protective behaviors. *Death toll exceeds 100k* represents the date the officially recorded COVID-19 death toll in the US exceeded 100,000 deaths (May 27, 2020). *CDC universal mask guidance* (July 2020) marks the US CDC’s recommendation that all individuals use face masks in indoor public settings. *Vaccine availability* (spring 2021) denotes the point at which COVID-19 vaccines became widely available to the US population without the eligibility restrictions imposed by the earlier prioritization strategies, reached for all adults in approximately April 2021. *Vaccination coverage 70%* (summer 2021) marks the point at which at least 70% of US adults had received one or more doses of a COVID-19 vaccine, reached in early August 2021. *Omicron dominant* (January 2022) marks the appearance and rapid rise to dominance of the Omicron (B.1.1.529) variant among US infections. *At-home tests widely available* (January 2022) marks the point at which rapid antigen self-tests became broadly accessible to the US public, coinciding with the launch of the federal free-distribution program and the requirement that insurers cover at-home tests. *Note: “going to a doctor or hospital” was assessed using the following survey item: “In the last 24 hours, did you or any members of your household do any of the following activities outside of your home? (Please select all that apply)”, with the relevant category being “Go to a doctor or visit a hospital”

**Fig 4.**
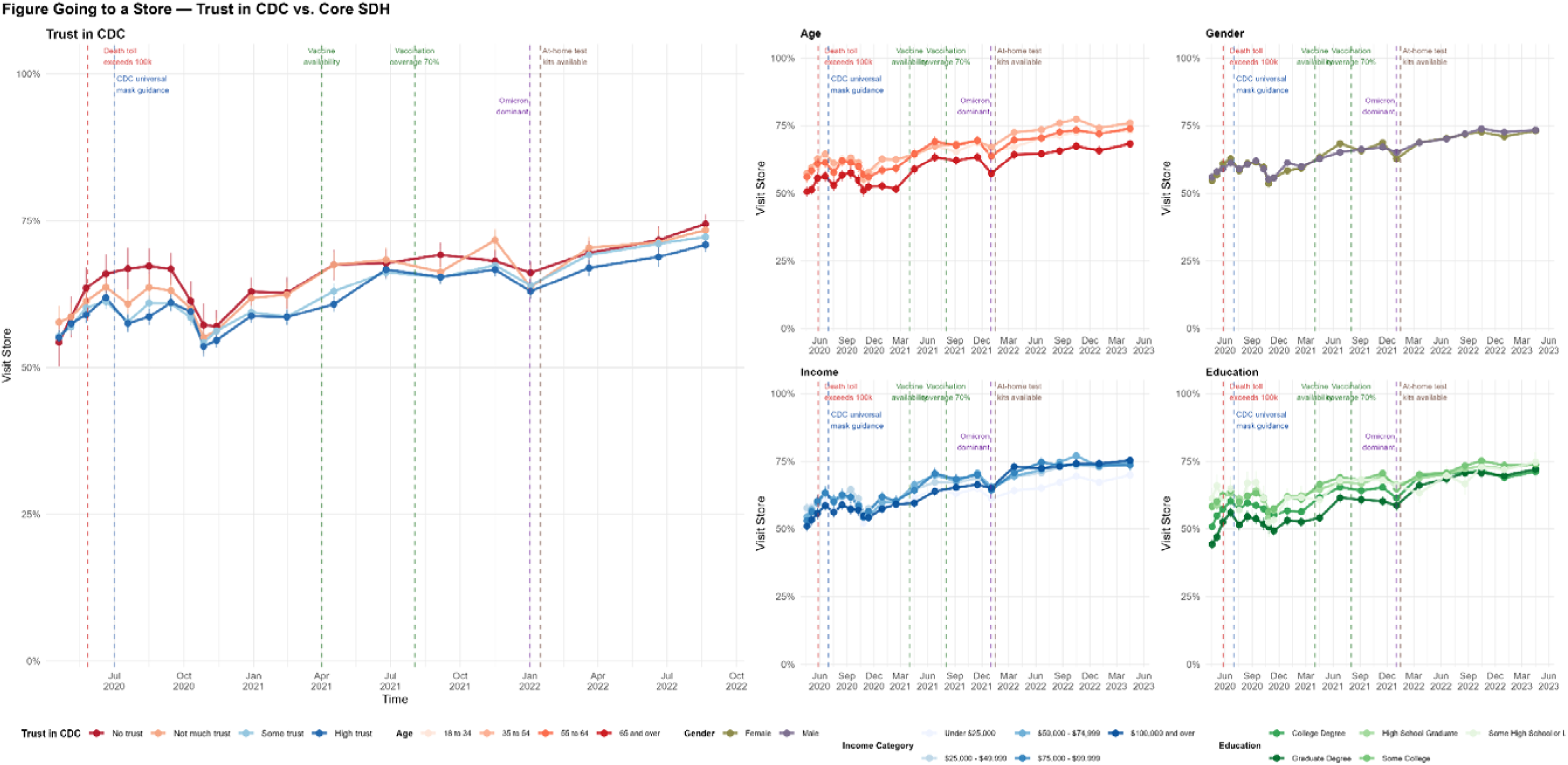
Temporal trajectory of going to a store, stratified by trust in the CDC and by core sociodemographic predictors (Age, Gender, Income, Education). The left panel shows the temporal evolution of the proportion of survey respondents reporting to have visited a store in the last 24 hours, stratified by self-reported trust in the CDC (high, some, not much, and none — corresponding to the survey response options “a lot”, “some”, “not too much”, and “not at all”). The four right-hand panels show the same outcome stratified by age, gender, household income, and educational attainment. Points represent survey-weighted estimates and vertical bars denote 95% confidence intervals, based on *20* waves of the CHIP50 Project survey. Dashed vertical lines mark key pandemic milestones with potential impact on protective behaviors. *Death toll exceeds 100k* represents the date the officially recorded COVID-19 death toll in the US exceeded 100,000 deaths (May 27, 2020). *CDC universal mask guidance* (July 2020) marks the US CDC’s recommendation that all individuals use face masks in indoor public settings. *Vaccine availability* (spring 2021) denotes the point at which COVID-19 vaccines became widely available to the US population without the eligibility restrictions imposed by the earlier prioritization strategies, reached for all adults in approximately April 2021. *Vaccination coverage 70%* (summer 2021) marks the point at which at least 70% of US adults had received one or more doses of a COVID-19 vaccine, reached in early August 2021. *Omicron dominant* (January 2022) marks the appearance and rapid rise to dominance of the Omicron (B.1.1.529) variant among US infections. *At-home tests widely available* (January 2022) marks the point at which rapid antigen self-tests became broadly accessible to the US public, coinciding with the launch of the federal free-distribution program and the requirement that insurers cover at-home tests. *Note: “going to a store” was assessed using the following survey item: “In the last 24 hours, did you or any members of your household do any of the following activities outside of your home? (Please select all that apply)”, with the relevant category being “Go to the store”

**Fig 5.**
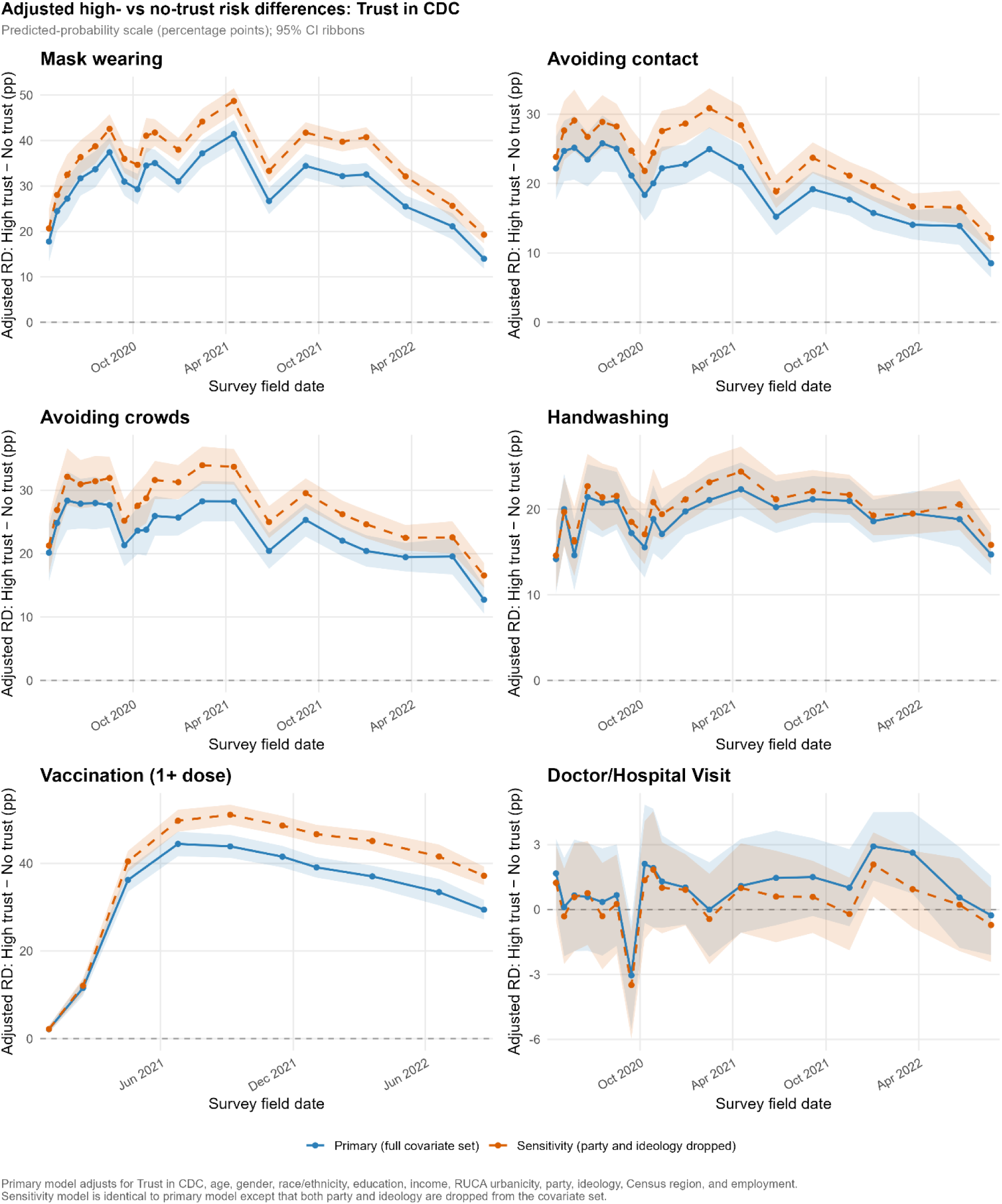
Covariate-adjusted risk differences in protective behaviors between high- and no-trust respondents (trust in the CDC), across survey waves. Each panel plots the adjusted risk difference (RD) in the probability of reporting a given behavior for respondents with high versus no trust in the CDC (High trust − No trust), estimated separately for each survey wave on the predicted-probability scale (percentage points). Estimates are derived from survey-weighted multivariable logistic regression models and expressed as differences in predicted probability; positive values indicate that high-trust respondents were more likely to report the behavior. Two specifications are shown for each outcome: the primary model (blue, solid) adjusts for trust in the CDC, age, gender, race/ethnicity, education, income, RUCA urbanicity, party, ideology, Census region, and employment, while the sensitivity model (orange, dashed) is identical except that party and ideology are omitted from the covariate set. Shaded ribbons denote 95% confidence intervals, and the dashed horizontal line marks a null risk difference of zero. Panels correspond to six behaviors — mask wearing, avoiding contact, avoiding crowds, handwashing, vaccination (≥1 dose), and visiting a doctor or hospital. Vertical axes are scaled independently across panels; in particular, Doctor/Hospital Visit is shown on a compressed scale that includes negative values, and the vaccination panel covers only the period from vaccine availability onward (i.e., after February 2021). Estimates are based on 20 waves of the CHIP50 Project survey.

We re-estimated the fully adjusted models substituting trust in other institutions for trust in the CDC. During the vaccine rollout (Wave 16), trust in doctors and hospitals produced an adjusted mask-wearing difference of similar magnitude to the CDC (34.7 pp, 95% CI 29.2–40.3, versus 37.2 pp for the CDC), and trust in state government produced an intermediate difference (21.8 pp, 19.4–24.2). Trust in banks was evaluated as a negative-control exposure and modeled jointly with trust in the CDC (the primary exposure) to examine shared confounding pathways.^25^ In marked contrast to trust in the CDC, the adjusted mask-wearing difference by trust in banks was small throughout the pandemic – 2.7 pp (95% CI 1.7 to 3.6) in the pre-vaccine period, 4.5 pp (3.4 to 5.7) during vaccine rollout, and 6.2 pp (4.7 to 7.7) during the Omicron period (**Figure 6, Supplementary Figures S11 – S16**). Within these jointly adjusted models, trust in the CDC remained a strong predictor of mask wearing, with high- versus no-trust adjusted odds ratios of 4.2 (95% CI 3.9–4.4) in the pre-vaccine period, 5.2 (4.9–5.5) during the vaccine rollout, and 3.2 (3.0–3.5) during the Omicron period (all p < 0.001, **Supplementary Figure S15**), underscoring that trust in the CDC remained the dominant behavioral predictor even in the negative control models. The association with vaccine uptake was close to the null (median 1.5 pp, vs. 36.6 pp for the CDC – an attenuation of approximately 96%) and across all behavioral outcomes the bank-trust differences were consistently a small fraction of the corresponding CDC differences (**Figure 5**).

**Fig 6.**
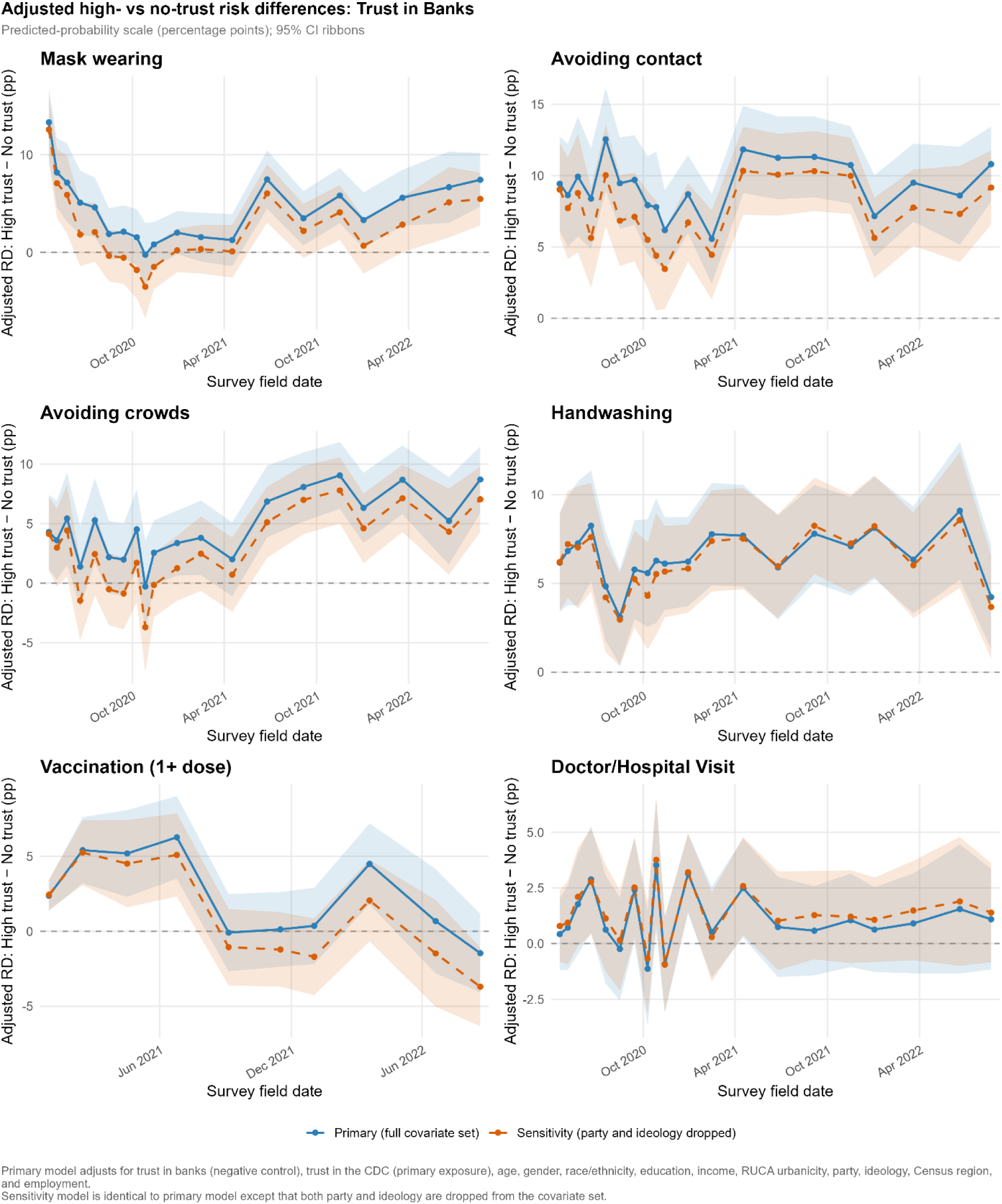
Covariate-adjusted risk differences in protective behaviors between high- and no-trust respondents (trust in banks; negative-control exposure), across survey waves. Each panel plots the adjusted risk difference (RD) in the probability of reporting a given behavior for respondents with high versus no trust in banks (High trust − No trust), estimated separately for each survey wave on the predicted-probability scale (percentage points). Trust in banks is included as a negative-control exposure — selected because it plausibly shares the general dispositional and sociodemographic confounders of trust in the CDC but has no direct causal pathway to protective behaviors. Estimates are derived from survey-weighted multivariable logistic regression models and expressed as differences in predicted probability; positive values indicate that high-trust respondents were more likely to report the behavior. Two specifications are shown for each outcome: the primary model (blue, solid) adjusts for trust in banks, trust in the CDC (the primary exposure), age, gender, race/ethnicity, education, income, RUCA urbanicity, party, ideology, Census region, and employment, while the sensitivity model (orange, dashed) is identical except that party and ideology are omitted from the covariate set. Shaded ribbons denote 95% confidence intervals, and the dashed horizontal line marks a null risk difference of zero. Panels correspond to six behaviors — mask wearing, avoiding contact, avoiding crowds, handwashing, vaccination (≥1 dose), and visiting a doctor or hospital. Vertical axes are scaled independently across panels; in particular, Mask wearing, Avoiding crowds, Vaccination, and Doctor/Hospital Visit are shown on a compressed scale that includes negative values, and the vaccination panel covers only the period from vaccine availability onward (i.e., after February 2021). Estimates are based on 20 waves of the CHIP50 Project survey.

These analyses are exploratory rather than aimed at causal inference; nonetheless, the specificity of the behavioral association to health- and government-relevant institutions and its marked attenuation for a non-health institution conditional on trust in the CDC is consistent with the differences reflecting institution-specific trust beyond a generic disposition to trust or distrust institutions. The bank-trust association was attenuated but not fully nullified, likely reflecting that trust in the CDC is one of several forms of trust that shape behavior and that trust in banks captures other dimensions of trust not fully reflected in trust in the CDC predictor.

Extending the adjusted models to two additional outcomes sharpened the behavioral interpretation of the trust contrast. Trust in the CDC was strongly associated with vaccination (Figure 5): as vaccines became broadly available, the adjusted high- versus no-trust gap in receipt of at least one dose widened to 44.4 pp (95% CI 41.6–47.2) at Wave 18 and remained substantial during the Omicron emergence in January 2022 at 39.1 pp (36.8-41.4)). In sharp contrast, the adjusted difference for visiting a doctor or hospital was negligible and non-significant during the first two years of the pandemic (0.6 pp [95% CI −1.9 to 3.1] pre-vaccine and 0.0 pp [−2.2 to 2.2] at rollout), with a weak association emerging during the Omicron period (2.9 pp [1.3 to 4.5]) that was an order of magnitude smaller than the corresponding differences in masking and vaccine uptake. Thus, after full adjustment, institutional trust is associated with large differences in voluntary protective behaviors — masking, distancing, and vaccine uptake — but essentially no difference in essential care-seeking, indicating that trust modifies discretionary, recommendation-driven behaviors rather than systemic healthcare seeking.

### Institutional trust contributes independent variance beyond political and social domains

To further quantify how much of the trust contrast is independent of the sociodemographic and political domains, we fit a hierarchical sequence of survey-weighted logistic regressions for each protective behavior and trust in the CDC, beginning with a trust-only baseline (M0) and sequentially adding demographic (M1), socioeconomic (M2), political (M3), and geographic (M4) covariate blocks (Table 1). For mask wearing, the adjusted high- versus no-trust difference was 40.9 pp in the trust-only model (median across 20 waves; interquartile range [IQR] 36.0–44.7) and was essentially unchanged by the addition of demographic and socioeconomic factors (39.1 pp in M1 and 37.8 pp in M2). The contrast attenuated appreciably when political identity — party identification and ideology — entered in M3, falling to 31.6 pp (IQR 26.7–34.2; a 16% reduction in the trust log-odds relative to baseline). Adding geographic context in M4 produced almost no further change (31.1 pp; IQR 26.5–33.7). Of the four covariate domains, only political identity shares appreciable explanatory power for the variance in the behavioral outcomes with institutional trust, and even after full adjustment roughly three quarters of the trust difference persist: in the fully adjusted model, respondents with high trust in the CDC were 31.1 pp more likely to always wear a mask than those with no trust (**Table 1**). The same ordering held for the other protective behaviors (**Supplementary Table S1**).

**Table 1.**
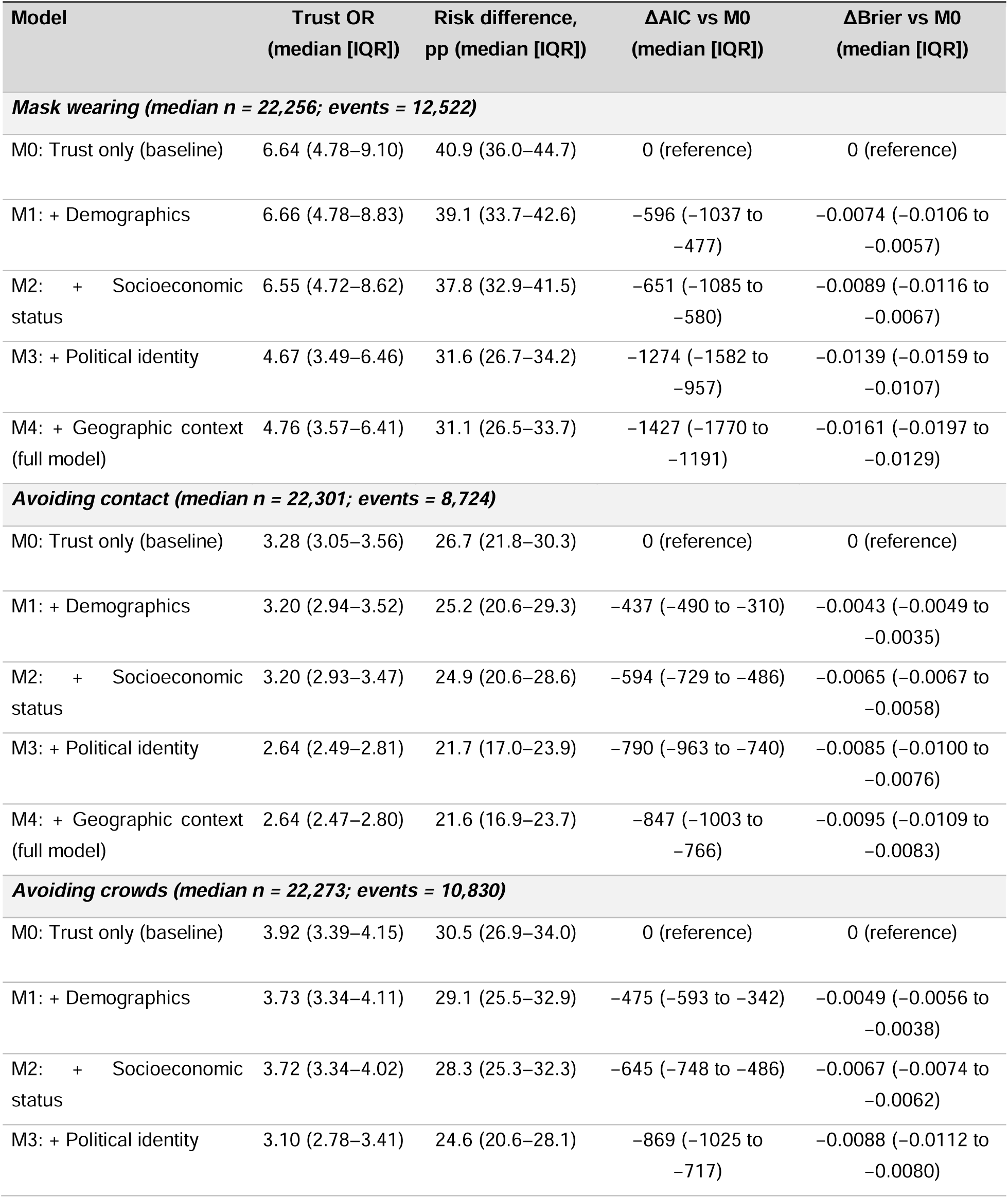
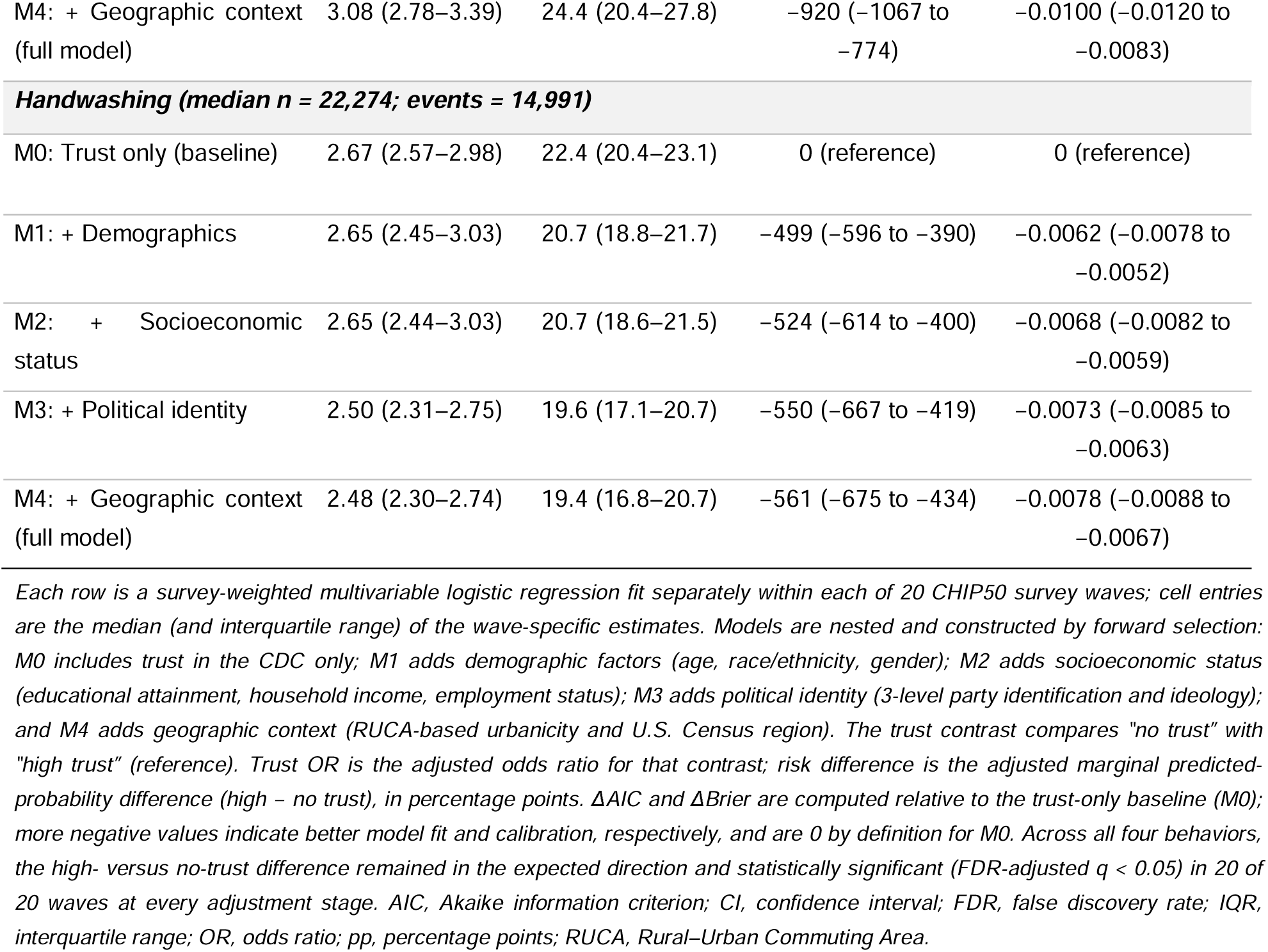
Sequential adjustment of the association between trust in the CDC and adoption of protective behaviors, CHIP50, 2020–2024. Each row is a survey-weighted multivariable logistic regression fit separately within each of 20 CHIP50 survey waves; cell entries are the median (and interquartile range) of the wave-specific estimates. Models are nested and constructed by forward selection: M0 includes trust in the CDC only; M1 adds demographic factors (age, race/ethnicity, gender); M2 adds socioeconomic status (educational attainment, household income, employment status); M3 adds political identity (3-level party identification and ideology); and M4 adds geographic context (RUCA-based urbanicity and U.S. Census region). The trust contrast compares “no trust” with “high trust” (reference). Trust OR is the adjusted odds ratio for that contrast; risk difference is the adjusted marginal predicted-probability difference (high − no trust), in percentage points. ΔAIC and ΔBrier are computed relative to the trust-only baseline (M0); more negative values indicate better model fit and calibration, respectively, and are 0 by definition for M0. Across all four behaviors, the high- versus no-trust difference remained in the expected direction and statistically significant (FDR-adjusted q < 0.05) in 20 of 20 waves at every adjustment stage. AIC, Akaike information criterion; CI, confidence interval; FDR, false discovery rate; IQR, interquartile range; OR, odds ratio; pp, percentage points; RUCA, Rural–Urban Commuting Area.

| Model | Trust OR<br>(median [IQR]) | Risk difference,<br>pp (median [IQR]) | ΔAIC vs M0<br>(median [IQR]) | ΔBrier vs M0<br>(median [IQR]) |
| --- | --- | --- | --- | --- |
| **Mask wearing (median n = 22,256; events = 12,522)** |  |  |  |  |
| M0: Trust only (baseline) | 6.64 (4.78–9.10) | 40.9 (36.0–44.7) | 0 (reference) | 0 (reference) |
| M1: + Demographics | 6.66 (4.78–8.83) | 39.1 (33.7–42.6) | –596 (–1037 to<br>–477) | –0.0074 (–0.0106 to<br>–0.0057) |
| M2: + Socioeconomic<br>status | 6.55 (4.72–8.62) | 37.8 (32.9–41.5) | –651 (–1085 to<br>–580) | –0.0089 (–0.0116 to<br>–0.0067) |
| M3: + Political identity | 4.67 (3.49–6.46) | 31.6 (26.7–34.2) | –1274 (–1582 to<br>–957) | –0.0139 (–0.0159 to<br>–0.0107) |
| M4: + Geographic context<br>(full model) | 4.76 (3.57–6.41) | 31.1 (26.5–33.7) | –1427 (–1770 to<br>–1191) | –0.0161 (–0.0197 to<br>–0.0129) |
| **Avoiding contact (median n = 22,301; events = 8,724)** |  |  |  |  |
| M0: Trust only (baseline) | 3.28 (3.05–3.56) | 26.7 (21.8–30.3) | 0 (reference) | 0 (reference) |
| M1: + Demographics | 3.20 (2.94–3.52) | 25.2 (20.6–29.3) | –437 (–490 to –310) | –0.0043 (–0.0049 to<br>–0.0035) |
| M2: + Socioeconomic<br>status | 3.20 (2.93–3.47) | 24.9 (20.6–28.6) | –594 (–729 to –486) | –0.0065 (–0.0067 to<br>–0.0058) |
| M3: + Political identity | 2.64 (2.49–2.81) | 21.7 (17.0–23.9) | –790 (–963 to –740) | –0.0085 (–0.0100 to<br>–0.0076) |
| M4: + Geographic context<br>(full model) | 2.64 (2.47–2.80) | 21.6 (16.9–23.7) | –847 (–1003 to<br>–766) | –0.0095 (–0.0109 to<br>–0.0083) |
| **Avoiding crowds (median n = 22,273; events = 10,830)** |  |  |  |  |
| M0: Trust only (baseline) | 3.92 (3.39–4.15) | 30.5 (26.9–34.0) | 0 (reference) | 0 (reference) |
| M1: + Demographics | 3.73 (3.34–4.11) | 29.1 (25.5–32.9) | –475 (–593 to –342) | –0.0049 (–0.0056 to<br>–0.0038) |
| M2: + Socioeconomic<br>status | 3.72 (3.34–4.02) | 28.3 (25.3–32.3) | –645 (–748 to –486) | –0.0067 (–0.0074 to<br>–0.0062) |
| M3: + Political identity | 3.10 (2.78–3.41) | 24.6 (20.6–28.1) | –869 (–1025 to<br>–717) | –0.0088 (–0.0112 to<br>–0.0080) |
| M4: + Geographic context (full model) | 3.08 (2.78–3.39) | 24.4 (20.4–27.8) | –920 (–1067 to –774) | –0.0100 (–0.0120 to –0.0083) |
| **Handwashing (median n = 22,274; events = 14,991)** |  |  |  |  |
| M0: Trust only (baseline) | 2.67 (2.57–2.98) | 22.4 (20.4–23.1) | 0 (reference) | 0 (reference) |
| M1: + Demographics | 2.65 (2.45–3.03) | 20.7 (18.8–21.7) | –499 (–596 to –390) | –0.0062 (–0.0078 to –0.0052) |
| M2: + Socioeconomic status | 2.65 (2.44–3.03) | 20.7 (18.6–21.5) | –524 (–614 to –400) | –0.0068 (–0.0082 to –0.0059) |
| M3: + Political identity | 2.50 (2.31–2.75) | 19.6 (17.1–20.7) | –550 (–667 to –419) | –0.0073 (–0.0085 to –0.0063) |
| M4: + Geographic context (full model) | 2.48 (2.30–2.74) | 19.4 (16.8–20.7) | –561 (–675 to –434) | –0.0078 (–0.0088 to –0.0067) |

Sequential adjustment improved model fit and probabilistic calibration: each successive covariate block lowered the Akaike Information Criterion (AIC) and reduced the survey-weighted Brier score relative to the trust-only baseline (the full masking model M4 improved on M0 by a median of roughly 1,400 AIC units, **Table 1**), indicating that the added domains carry a substantial amount of non-redundant behavioral information. However, in a leave-one-block-out analysis of the fully adjusted model, removing trust degraded fit more than removing any other covariate block and roughly twice as much as removing the political block (median ΔAIC +921 [IQR 545–1181] for trust versus +442 [361–581] for party and ideology; median increase in the survey-weighted Brier score +0.0096 versus +0.0041), while demographics (+368), geography (+180), and socioeconomic status (+33) contributed less (**Supplementary Table S1**). This ranking was most consistent for masking and social-distancing behaviors, whereas for handwashing and during the Omicron era demographic factors contributed comparable or greater model fit improvements.

### Trust differences are behaviorally specific: trust affects voluntary compliance but not healthcare seeking

When trust-related adherence differences were examined across the full behavioral spectrum, we found that trust in the CDC was significantly associated with the four primary protective behaviors in 100% of waves but showed essentially no association with visiting a doctor or hospital. This relationship was significant in only 1 of 20 waves (**Figure 3**), with pairwise risk differences below 2 pp throughout all pandemic periods, an order of magnitude smaller than the gaps for masking. Intermediate associations were observed for going to a store (**Figure 4**, significant in 50% of waves) and going to work (75%), consistent with these activities reflecting a mix of discretionary choice and structural necessity. Healthcare-seeking behavior was instead governed by access-related determinants: income, education, and race/ethnicity showed significant associations with doctor and hospital visits in 87%, 91%, and 74% of waves, respectively. This behavioral specificity — trust primarily governs voluntary compliance with recommendations but not healthcare-seeking — has direct consequences for model parameterization, as we discuss below.

### Declines in institutional trust mirror declines in protective behaviors

The temporal structure of behavior change during the pandemic further provides a case for trust as a dynamic model parameter. Trust changes while other variables such as demographics do not or barely do. Previous studies^26^ decomposed population-level protective behaviors into two components: a declining linear trend and an oscillatory response synchronized with officially reported COVID-19 mortality. Social distancing (avoiding contacts and crowds) declined approximately threefold between April 2020 and May 2022 — a decrease that cannot be fully explained by feedback from mortality, which rebounded repeatedly over the same period. The 31-pp erosion of high trust documented by Perlis et al. (2024) over the course of the pandemic (April 2020-January 2024) provides one plausible mechanism for this decline: as institutional confidence fell, the perceived legitimacy of sustained protective behavior may have declined in parallel. Consistent with this interpretation, the behavioral decline was non-uniform across the political landscape — participants in Democratic-leaning states reported higher baseline adoption of protective behaviors than their Republican-leaning counterparts even at equivalent levels of policy stringency^26^ mirroring the trust differences that our analysis identifies as independent predictors of behavior. Correspondingly, the magnitude of the adjusted trust differences in protective behaviors was largest during the pre-vaccine period and vaccine rollout — when institutional guidance was most directive and trust most consequential for behavioral choices — and narrowed during the Omicron era as both trust levels and overall protective behavior adoption declined across the population (**Figures 1, 2, and 5**). This narrowing of the trust gap may be attributed to decreased clarity in public health messaging, shifts in policy attitudes away from absolute prevention toward mitigation focused on high-risk populations, or broad changes in population attitudes toward protective behaviors (i.e., pandemic fatigue), clearer evidence about which sectors of the population were at higher risk, and the advent of pharmaceutical interventions later in the pandemic.

## Discussion

Our results highlight institutional trust as a dominant, temporally stable predictor of reported adoption of protective behaviors during the COVID-19 pandemic, exceeding age, income, educational attainment, and partisan affiliation across a variety of risk-preventing behaviors during the years of strongest pandemic activity. The four- to six-fold increased adherence gap by trust compared to conventional sociodemographic variables suggests that using demographic characteristics instead of constructs like institutional trust can lead to underestimation of behavioral variability. Critically, our sequential comparisons of adjusted models move this claim beyond a contrast of unadjusted differences and highlight that institutional trust is not merely a proxy for political affiliation. Although institutional trust shares explanatory variance with partisanship — the only covariate domain whose inclusion meaningfully attenuated the trust difference — trust remains an independent and structurally relevant predictor of protective behavior. In the fully adjusted model, removing trust degraded model fit more than removing any other covariate block, incurring an information loss (AIC penalty) roughly twice that of removing partisanship and far exceeding that of sociodemographic or geographic covariates. Conventional modeling approaches that stratify transmission heterogeneity (assortativity) by demographic factors alone are therefore systematically omitting behavioral variability driving transmission.

A model that assigns the same contact-reduction rate to low- and high-trust individuals within the same sociodemographic group will, during periods of intensified public health interventions, overestimate effective contacts among high-trust individuals and underestimate them among low-trust individuals. These errors would likely accumulate and compound over protracted epidemics. The evidence from our model comparisons makes this concrete: just as omitting trust sharply degraded the probabilistic calibration of our multivariable regressions (the largest increase in survey-weighted Brier score of any covariate block), omitting trust from population-level transmission models is likely to introduce substantial parameterization error in the effective contact rate. Such errors would be greatest during periods of rapid trust erosion, as observed throughout the COVID-19 pandemic and during other large-scale outbreaks such as the 2013–2016 West African Ebola epidemic, in which collapsing community confidence in the public health response materially shaped transmission and the success of control efforts^27–30^.

This argument is sharpened by two forms of specificity indicating that institutional trust represents a genuine epidemiological parameter rather than a generalized trait. Behaviorally, trust predicted whether people reported to wear masks, avoid crowds, wash their hands, and get vaccinated, but showed essentially no association with whether they visited a doctor or hospital; the gap in care-seeking by trust neared zero after adjustment. Institutionally, the adherence gap by trust narrowed by approximately 85% when trust in banks was substituted as a negative control, whereas trust in doctors and hospitals and in state government retained larger gaps. In conjunction, this dual specificity indicates that institutional trust captures behaviorally and institutionally specific dimensions of behavioral adaptation impacting disease transmission. For epidemic models, this implies that trust should be incorporated as a modifier specific to voluntary contact-reduction and vaccination rates, whereas care-seeking behavior should be parameterized separately; conflating these two pathways produces a different class of error than omitting trust entirely. Therefore, integration of trust into any modeling efforts should consider these two types of behaviors differently as for a single level of trust, individuals can demonstrate variation in their engagement with the different types of protective behaviors. One possible mechanism for this variation can be viewed through the lens of the Health Belief Model^14,15,31^: those with low trust in health institutions may not view proposed restrictions as signals of increased infection risk and therefore do not modulate their behavior unless absolutely essential; this lack of adaptation only resolves once medical consequences become serious or life-threatening.

The linear behavioral decline in protective behaviors documented previously^26^ emphasizes our rationale for modeling trust as a dynamic state variable. Standard epidemiological approaches to waning immunity in epidemiological frameworks offer a useful conceptual analogy: just as individuals transition from recovered to susceptible compartments at a rate governed by immunological decay^32,33^, population members can transition from high- to low-trust strata at a rate governed by epidemic trajectory, institutional communication quality, and social contagion of distrust via information networks. Additionally, the approximately linear decline in trust documented previously^12^ provides initial evidence for the functional form of this transition, though whether it reflects a continuous process or step changes triggered by specific communication or policy failures remains an open question that longitudinal survey data can help resolve. Despite clear evidence for this linear decline, we caution the community against attributing the decline to solely degradations in institutional trust that spread to other types of human behavior during the pandemic, as it is understood that engagement in protective behaviors decreased throughout the pandemic as “pandemic fatigue” affected many individuals who were subject to preventive restrictions^34^.

Besides this linear decline in behaviors, we find little evidence of responses in protective behaviors to discrete policy actions or public health communication (key pandemic milestones were visually annotated in Figures 1 – 3). Except for the vaccine-rollout period from early to mid-2021, when masking fell steeply as vaccines became available and the CDC relaxed guidance for vaccinated individuals, protective behaviors showed no abrupt changes linked to other milestones, such as the Omicron emergence. Behaviors instead broadly tracked the gradual decline with some oscillations linked to mortality, reinforcing our argument that behavioral adaptation was governed less by discrete announcements than by the slow erosion of institutional trust and the continuing pandemic threat.

Building on conceptual work on risk-behavior stratification^35^, we propose dividing the population into strata by institutional trust—each sized proportionately to survey responses and assigned distinct contact and vaccination-uptake rates. During active public health guidance, low-trust individuals are less likely to mask, avoid crowds, or distance, and therefore expected to make more transmission-relevant contacts than high-trust individuals. For example, choosing three compartments, we denote the group-specific transmission rates as {β*_h_*, {β_m_, and {β_l_ (for high, moderate, and low trust), and assume {β*_l_* > {β*_m_* > {β*_h_* — that is, transmission risk is highest among those with the least institutional trust. Similarly, vaccine uptake rates v*_h_*, v*_m_* , v*_l_* differ across trust strata: the findings of Perlis et al.^12^ imply v*_h_*» v*_i_* , with high-trust individuals nearly five times more likely to be vaccinated than low-trust individuals; this aligns with existing work emphasizing the direct connection between trust and vaccine acceptance^36–38^. A key practical advantage of this structure is that both the transmission rate differentials and the vaccination rate differentials can be estimated from survey-reported behaviors and linked to transmission dynamics through calibration. This matters because inferring behavioral parameters from the epidemic data they are meant to explain is inherently circular and cannot distinguish between different behavioral mechanisms.

Beyond the static stratification, the model should capture two distinct behavioral dynamics as evidenced by our data. The first is a long-term decline: protective behaviors decreased steadily over the course of the pandemic regardless of epidemic severity, consistent with eroding institutional trust. The second is a short-term oscillation: protective behaviors rose when COVID-19 deaths were high and fell when deaths were low. We represent the total contact-reduction rate for a trust stratum r at time t as the sum of these two components: *C(t, τ) = B(t, τ) + φ(M(t), τ)* where B captures the decline and rp captures the mortality-driven oscillation. The overall transmission rate for that stratum is then *β(t, τ) = {β_0_(τ) ·* [1 *- C(t, τ)*], where *β_0_(τ)* is the baseline transmission rate in absence of any behavioral modification. To include feedback between disease severity and behaviors, the oscillatory term φ should increase with rising mortality (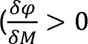, meaning people become more cautious as deaths rise^26^). Together, these components provide an empirically grounded extension of the standard SEIR framework — one whose parameters can be estimated from the same longitudinal survey infrastructure that generated the findings reported here.

We anticipate the framework will need to consider intricacies related to temporal dynamics of infectious diseases. Low-trust individuals getting infected before high-trust individuals (in addition to being infected at different/higher rates) would lead to several implications. For example, if the trust differences between individuals (i.e., the relative change between high-trust and low-trust individuals) is temporally stable (our data, as seen in **Supplementary Figure S19**, shows this stability), then a turnover in relative incidence of the infection due to saturation of susceptibility among the low trust group could occur^39^. This leads to changes in the critical immunity fraction, which would go down when there are differences between the trust groups and go back up when the trust groups homogenize^40^. This relationship is akin to previously evaluated temporal phenomena^10^ where less densely populated US counties had lower COVID-19 mortality during spring but higher mortality in the winter due to changes in the underlying population immunity.

The implications extend beyond model structure to surveillance design and intervention strategies. Prospectively, near-real-time trust monitoring should be integrated with epidemiological surveillance — analogous to utilizing wastewater monitoring to provide early warning of pathogen spread^41^. Researchers would gain the ability to not only understand how trust in institutions varies in periods outside of crises (i.e., in the absence of an epidemic) but also how the onsets of such crises are affected by differential trust across population groups. Like regular polls about politically relevant themes that are conducted weekly or monthly, measures of trust (in both health and non-health institutions) can provide valuable insights to both public health authorities and other decision makers. Our data help to identify the populations for whom this monitoring is most urgent: those with historically low baseline trust, including rural communities, younger adults, and certain racial and ethnic minorities^12,42–45^ as well as groups in whom trust eroded most rapidly during the COVID-19 pandemic. Default trust-stratified behavioral parameters derived from the COVID-19 experience should be developed for use in novel mathematical models, analogous to the role age-stratified contact matrices from studies like POLYMOD^5^ have played for decades.

Several limitations warrant acknowledgment. Although our fully adjusted model (M4) provides a robust descriptive characterization, the covariates it conditions on are not interchangeable statistical controls: political identity may operate as a mediator of the trust–behavior relationship, or as a competing predictor or collider, rather than as a conventional confounder. The estimates therefore represent adjusted descriptive associations rather than causal effects, given the observational design—though the lagged analyses in Perlis et al. (2024) are consistent with trust preceding vaccination. Likewise, the parallel decline in trust and protective behaviors is consistent with a trust-mediated mechanism but cannot be separated from pandemic fatigue□□-□□, which could produce a similar temporal pattern independently of trust. Disentangling these processes would require within-person longitudinal designs in which individual-level changes in trust are linked to concurrent changes in behavior, controlling for time elapsed since the pandemic onset. Trust in individual institutions was measured as a single survey item, which cannot capture the multidimensional nature of institutional trust; validated multi-item scales should be incorporated into future surveys^46,47^. Furthermore, protective behavior differences were estimated from survey-reported behavioral indicators and parameterization will require linkage to changes in transmission intensity through model calibration.

Our negative-control-exposure framework²□ probes the plausibility of an institution-specific interpretation and gauges unmeasured confounding rather than supporting formal causal claims. Trust in banks benchmarks the association for trust in the CDC but is an imperfect negative control□¹: conditional on trust in the CDC, the bank association was attenuated but not fully nullified. This residual is expected—trust in the CDC is only one form of institutional trust, and trust in banks likely captures a broader dispositional or anti-institutional orientation that the CDC measure does not absorb. Critically, our interpretation rests on the pronounced difference in magnitude between the two exposures: across all outcomes, the adjusted behavioral differences for trust in the CDC exceeded those for trust in banks by a wide margin — most strikingly for mask wearing and vaccine uptake by an order of magnitude. This specificity provides strong, if informal, evidence that the observed behavioral differences are driven by trust in public-health institutions, rather than by a generic anti-establishment skepticism or a population-wide disposition to distrust. The appropriate functional forms for trust-behavior coupling in compartmental models remain empirically undetermined with linear, threshold, and nonlinear specifications implying different epidemic dynamics. Finally, the evidence is restricted to survey data collected in the US; trust differences may differ across countries and temporal contexts, though analogous trust heterogeneity in vaccine hesitancy has been documented internationally^48^. Despite extensive validation against probability designs^49–51^ CHIP50 is a nonprobability sample, which limits representativeness for small subgroups and low-trust individuals may be underrepresented, potentially attenuating estimated trust gaps.

These limitations notwithstanding, the implications of our findings are clear. The COVID-19 pandemic has generated an unusually rich and temporally resolved record of how population-level behavior changes during a sustained public health crisis — and that record delivers a consistent message: institutional trust is a critical social determinant of whether individuals adopt and sustain protective behaviors. As a predictor, trust exceeds age, income, education, and partisan affiliation in explanatory magnitude, and, crucially, this advantage persists under rigorous multivariable adjustment: trust contributes independent structural variance that is not reducible to political or sociodemographic differences. It is also not static: trust in medical institutions eroded by more than 31 percentage points over four years, shifting the behavioral landscape in ways that no model with fixed demographic parameters can capture. Furthermore, its effects reflect two dimensions of specificity: how behaviors like mask-wearing were associated with institutional trust but seeking medical care was not, and how trust in different institutions (i.e., CDC vs. banks) were tied to fundamentally different behavioral outcomes.

Mathematical models that omit this variable are working with a structurally incomplete representation of the populations they describe. Demographically-stratified contact matrices capture an important dimension of heterogeneity, but they rarely approximate the proximate social and psychological forces that determine whether the individuals within any group will adopt the behaviors those models assume. The data infrastructure needed to close this gap already exists: the CHIP50 surveys have been shown to track COVID-19 infection rates longitudinally at national and state levels^52^ , and the behavioral data derived from the same surveys correlate strongly with COVID-19 mortality and hospitalization across pandemic waves^26^. The methodological path from measurement to model parameterization is clear.

What remains is to treat institutional trust as a primary model parameter that drives independent variance in protective behaviors beyond political and sociodemographic divides. Trust should thus be measured prospectively alongside epidemiological indicators, incorporated structurally using the framework proposed here, and updated in real time as future outbreaks unfold. The hidden parameters driving epidemic dynamics are not only biological. They are social, measurable, and they have been hiding in plain sight.

## Methods

### Survey design and data collection

We analyzed data from CHIP50^24^, a nationally representative nonprobability online survey conducted approximately every one to two months from April 2020 through January 2024. The survey was developed and overseen by a consortium of academic sites formed early in the pandemic to understand COVID-19–related attitudes and behaviors. The full survey design, institutional review board approval (Harvard University, exempt), and data collection procedures are described in detail elsewhere^12^. Participants provided informed consent online. The following analyses cover waves 1 – 24 (waves 4, 6, 8, and 15 were excluded due to insufficient sample size for the analyzed variables). Surveys used recruitment quotas for age, gender, and race/ethnicity within each state. Responses were weighted using interlocking national weights for age, gender, race/ethnicity, educational level, and region based on 2019 US Census data. This approach has been validated against probability-sampled designs and administrative data^49,50^.

### Behavioral outcomes and stratifiers

The primary outcomes were always wearing a mask outside the home, always avoiding contact with others, always avoiding crowded places, and frequent handwashing, each operationalized as binary outcome by dichotomizing survey responses at the “always” or “frequent” threshold consistent with prior analyses of these data^26^. We additionally examined routine and essential activities, such as visiting a store or visiting a doctor or hospital — to assess whether trust effects were specific to recommended protective behaviors. Vaccination status (receipt of at least one dose of a COVID-19 vaccine) was examined as a secondary outcome.

Analyzed institutional predictors were trust in the CDC, trust in physicians and hospitals, trust in state governments, and trust in banks, each measured by asking, “How much do you trust the following people and organizations to do what is right?” with four response options: a lot, some, not too much, or not at all. In waves prior to August 2022, a functionally equivalent variant was used; responses to the two versions were highly correlated within individuals (Spearman ρ = 0.76)^12^ and were treated as equivalent. Trust in CDC, trust in physicians and hospitals, trust in state governments, and trust in banks were analyzed as separate constructs throughout.

### Statistical analysis

Adherence to protective behaviors, the percentage of respondents reporting to follow a certain behavior, was stratified by trust in the CDC, coded as a four-level ordered factor with “no trust” as the reference category, partisan leaning, ideology, age groups, gender, race/ethnicity, household income, educational attainment, employment status, urbanicity (defined using Rural–Urban Commuting Area [RUCA] codes), and region. These were selected to represent the full range of sociodemographic, political, and institutional predictors of protective behavior identified in prior literature, enabling direct comparison of the magnitude of trust-related behavioral differences against conventional predictors. Secondary analyses substituted trust in doctors and hospitals, state governments, and banks for trust in the CDC; trust in banks was evaluated as a negative-control exposure to detect the potential impact of confounding^25^ assuming that trust in financial institutions should not predict the adoption of pandemic protective behaviors.

### Descriptive analyses

Within each survey wave, between-group differences in adherence were estimated as pairwise risk differences using survey-weighted generalized linear models with Taylor-series linearization for variance estimation. The statistical significance of each association between behavioral outcome and stratifier was assessed using omnibus design-based Wald F-tests; p-values were adjusted using the Benjamini-Hochberg false discovery rate (FDR) correction to correct for multiple testing across waves. To characterize temporal consistency, we summarized the proportion of waves in which each stratifier showed a statistically significant association after FDR correction. To provide concise comparisons, we highlight analyses conducted at representative landmark waves during three pandemic eras — pre-vaccine (wave 5), vaccine rollout (wave 16), and Omicron dominance (wave 21).

### Multivariable adjusted models

For each of the protective behaviors — always wearing a mask, always avoiding close contact, always avoiding crowds, and frequent handwashing — we fit a survey-weighted multivariable logistic regression separately for each of the CHIP50 waves. All models were estimated using post-stratification survey weights, with standard errors obtained via Taylor-series linearization. The primary analyses examined trust in the CDC and adjusted simultaneously for age, race/ethnicity, educational attainment, household income, urbanicity), partisan leaning (Independent as the reference), ideology (liberal, moderate, conservative; with moderate as the reference), U.S. Census region, and employment status. Adjusted behavior differences between high- versus low-trust groups were computed from the model estimates as marginal predicted-probability contrasts, standardized over the observed joint distribution of sociodemographic covariates.

Secondary analyses were conducted analogously for trust in doctors and hospitals and trust in state governments, using the same sociodemographic adjustment set. Negative-control models using trust in banks were further adjusted for trust in the CDC, so that the negative-control exposure was evaluated conditional on the primary exposure to detect potential residual confounding.^25^

To assess whether the behavioral differences by trust were confounded by partisanship, we conducted sensitivity analyses in which party leaning and ideology were removed from the covariate set; comparison of the trust contrasts across the two model specifications quantifies the extent to which partisan identity may mask the trust difference in behaviors. In a separate set of sensitivity analyses, models for the primary trust predictors (CDC, doctors/hospitals, and state governments) were additionally adjusted for a continuous index of general trust in others. This allowed us to isolate the behavioral influence of institution-specific trust from a baseline dispositional propensity toward interpersonal trust.

### Sequential adjustment and model fit

To characterize the independent explanatory contribution of institutional trust to protective behaviors, we evaluated a hierarchical, forward-selection sequence of nested survey-weighted logistic regressions. The sequence proceeded from a trust-only baseline model (M0; protective behavior regressed on trust in the CDC) through the stepwise addition of four covariate blocks: demographic characteristics (age, race/ethnicity, gender; M1), socioeconomic status (educational attainment, household income, employment status; M2), political identity (party leaning, ideology; M3), and geographic context (urbanicity and U.S. Census region; M4). Tracking the adjusted high- versus low-trust contrast across this hierarchy quantified the attenuation of the trust effect and thereby identified the covariate domains that share variance with — and therefore partially account for — the trust contrast. Relative model fit and parsimony were evaluated using the Akaike Information Criterion (AIC), with ΔAIC computed both against the trust-only baseline M0 and against the immediately preceding model in the sequence to quantify the incremental gain in information loss attributable to each covariate block. Overall model performance and probabilistic calibration were assessed using survey-weighted Brier scores, which provide a design-consistent measure of the mean squared deviation between model-implied probabilities and observed binary outcomes. To isolate the explanatory contribution of the trust and partisan blocks once all remaining domains were conditioned on, we performed a leave-one-block-out analysis on the fully adjusted model M4: each covariate block was omitted in turn and the resulting changes in the trust contrast, the AIC, and the Brier score were compared with those of the full model.

All analyses were conducted in R version 4.5.1 (2025-06-13 ucrt, “Great Square Root”).

## Data Availability

Anonymized numeric and ordinal survey data from the CHIP50 survey are deposited at https://github.com/MIGHTE_lab/trust_pandemic_driver and will be made publicly available upon journal acceptance.

## Data Availability

Additional data supporting this study are available from the corresponding author upon reasonable request.

## Code Availability

Analytic scripts and files (in R) used to process and analyze the CHIP50 survey data are deposited at https://github.com/MIGHTE_lab/trust_pandemic_driver_and will be made publicly available upon journal acceptance.

## Acknowledgments

This work was supported by CDC (CDC-RFA-FT-23-0069), NSF (SES-2029292, SES-2029792, SES-2116465, SES-2116189, SES-2116458, SES-211663, SES-2241884, SES-2241885, SES-2241886, SES-2241887), the Knight Foundation, Amazon Web Services, and the Peterson Foundation. AI language model assistance Claude (claude-sonnet-4-6, Anthropic) was used in editing and proofreading the manuscript text; all intellectual content and scientific claims were developed and verified by the authors.

## Author Contributions

Conceptualization: GD, AZ, MS

Data Collection: AU, KO

Formal analysis: GD, AZ

Funding acquisition: MS, KO, MB, JD, RP, DL

Investigation: GD, AZ, MS

Methodology: GD, AZ, MS, ML

Software: GD, AZ

Supervision: MS, KO, MB, JD, RP, DL

Writing - original draft: GD, AZ, MS

Writing - review and editing: GD, AZ, MS, KO, WH, ML, MB, AU, JD, RP, DL

## Competing Interests

R.P. reported receiving funding for consulting or service on scientific advisory boards from Vault Health, Belle Artificial Intelligence, Swan AI Studios, Mila Health, Alkermes, Genomind, Takeda, Circular Genomics, and Psy Therapeutics; holding equity in Circular Genomics, Psy Therapeutics, and Vault Health. M.S. reported receiving institutional research funds from the Johnson & Johnson Foundation, Janssen Global Public Health, and Pfizer Pharmaceuticals. No other competing interests declared.

---

**Fig S1 | Temporal trajectory of mask wearing, stratified by trust in the CDC and by secondary sociodemographic predictors (Political Affiliation, Race/Ethnicity, Employment status, Urbanicity).**

The left panel shows the temporal evolution of the proportion of survey respondents reporting always wearing a mask in public, stratified by self-reported trust in the CDC (high, some, not much, and none — corresponding to the survey response options “a lot”, “some”, “not too much”, and “not at all”). The four right-hand panels show the same outcome stratified by political affiliation, race/ethnicity, employment status, and urbanicity. Points represent survey-weighted estimates and vertical bars denote 95% confidence intervals, based on *20* waves of the CHIP50 Project survey. Dashed vertical lines mark key pandemic milestones with potential impact on protective behaviors. *Death toll exceeds 100k* represents the date the officially recorded COVID-19 death toll in the US exceeded 100,000 deaths (May 27, 2020). *CDC universal mask guidance* (July 2020) marks the US CDC’s recommendation that all individuals use face masks in indoor public settings. *Vaccine availability* (spring 2021) denotes the point at which COVID-19 vaccines became widely available to the US population without the eligibility restrictions imposed by the earlier prioritization strategies, reached for all adults in approximately April 2021. *Vaccination coverage 70%* (summer 2021) marks the point at which at least 70% of US adults had received one or more doses of a COVID-19 vaccine, reached in early August 2021. *Omicron dominant* (December 2021) marks the appearance and rapid rise to dominance of the Omicron (B.1.1.529) variant among US infections. *At-home tests widely available* (January 2022) marks the point at which rapid antigen self-tests became broadly accessible to the US public, coinciding with the launch of the federal free-distribution program and the requirement that insurers cover at-home tests.

**Fig S2 | Temporal trajectory of handwashing, stratified by trust in the CDC and by secondary sociodemographic predictors (Political Affiliation, Race/Ethnicity, Employment status, Urbanicity).**

The left panel shows the temporal evolution of the proportion of survey respondents reporting always washing their hands, stratified by self-reported trust in the CDC (high, some, not much, and none — corresponding to the survey response options “a lot”, “some”, “not too much”, and “not at all”). The four right-hand panels show the same outcome stratified by political affiliation, race/ethnicity, employment status, and urbanicity. Points represent survey-weighted estimates and vertical bars denote 95% confidence intervals, based on *20* waves of the CHIP50 Project survey. Dashed vertical lines mark key pandemic milestones with potential impact on protective behaviors. *Death toll exceeds 100k* represents the date the officially recorded COVID-19 death toll in the US exceeded 100,000 deaths (May 27, 2020). *CDC universal mask guidance* (July 2020) marks the US CDC’s recommendation that all individuals use face masks in indoor public settings. *Vaccine availability* (spring 2021) denotes the point at which COVID-19 vaccines became widely available to the US population without the eligibility restrictions imposed by the earlier prioritization strategies, reached for all adults in approximately April 2021. *Vaccination coverage 70%* (summer 2021) marks the point at which at least 70% of US adults had received one or more doses of a COVID-19 vaccine, reached in early August 2021. *Omicron dominant* (December 2021) marks the appearance and rapid rise to dominance of the Omicron (B.1.1.529) variant among US infections. *At-home tests widely available* (January 2022) marks the point at which rapid antigen self-tests became broadly accessible to the US public, coinciding with the launch of the federal free-distribution program and the requirement that insurers cover at-home tests.

**Fig S3 | Temporal trajectory of mask wearing, stratified by trust in banks (negative control exposure) and by core sociodemographic predictors (Age, Gender, Income, Education).** The left panel shows the temporal evolution of the proportion of survey respondents reporting always wearing a mask in public, stratified by self-reported trust in banks (high, some, not much, and none — corresponding to the survey response options “a lot”, “some”, “not too much”, and “not at all”). The four right-hand panels show the same outcome stratified by age, gender, household income, and educational attainment. Points represent survey-weighted estimates and vertical bars denote 95% confidence intervals, based on *20* waves of the CHIP50 Project survey. Dashed vertical lines mark key pandemic milestones with potential impact on protective behaviors. *Death toll exceeds 100k* represents the date the officially recorded COVID-19 death toll in the US exceeded 100,000 deaths (May 27, 2020). *CDC universal mask guidance* (July 2020) marks the US CDC’s recommendation that all individuals use face masks in indoor public settings. *Vaccine availability* (spring 2021) denotes the point at which COVID-19 vaccines became widely available to the US population without the eligibility restrictions imposed by the earlier prioritization strategies, reached for all adults in approximately April 2021. *Vaccination coverage 70%* (summer 2021) marks the point at which at least 70% of US adults had received one or more doses of a COVID-19 vaccine, reached in early August 2021. *Omicron dominant* (December 2021) marks the appearance and rapid rise to dominance of the Omicron (B.1.1.529) variant among US infections. *At-home tests widely available* (January 2022) marks the point at which rapid antigen self-tests became broadly accessible to the US public, coinciding with the launch of the federal free-distribution program and the requirement that insurers cover at-home tests.

**Fig S4 | Temporal trajectory of mask wearing, stratified by trust in banks (negative control exposure) and by secondary sociodemographic predictors (Political Affiliation, Race/Ethnicity, Employment status, Urbanicity).**

The left panel shows the temporal evolution of the proportion of survey respondents reporting always wearing a mask in public, stratified by self-reported trust in banks (high, some, not much, and none — corresponding to the survey response options “a lot”, “some”, “not too much”, and “not at all”). The four right-hand panels show the same outcome stratified by political affiliation, race/ethnicity, employment status, urbanicity. Points represent survey-weighted estimates and vertical bars denote 95% confidence intervals, based on *20* waves of the CHIP50 Project survey. Dashed vertical lines mark key pandemic milestones with potential impact on protective behaviors. *Death toll exceeds 100k* represents the date the officially recorded COVID-19 death toll in the US exceeded 100,000 deaths (May 27, 2020). *CDC universal mask guidance* (July 2020) marks the US CDC’s recommendation that all individuals use face masks in indoor public settings. *Vaccine availability* (spring 2021) denotes the point at which COVID-19 vaccines became widely available to the US population without the eligibility restrictions imposed by the earlier prioritization strategies, reached for all adults in approximately April 2021. *Vaccination coverage 70%* (summer 2021) marks the point at which at least 70% of US adults had received one or more doses of a COVID-19 vaccine, reached in early August 2021. *Omicron dominant* (December 2021) marks the appearance and rapid rise to dominance of the Omicron (B.1.1.529) variant among US infections. *At-home tests widely available* (January 2022) marks the point at which rapid antigen self-tests became broadly accessible to the US public, coinciding with the launch of the federal free-distribution program and the requirement that insurers cover at-home tests.

**Fig S5 | Temporal trajectory of avoiding contacts, stratified by trust in the CDC and by core sociodemographic predictors (Age, Gender, Income, Education).**

The left panel shows the temporal evolution of the proportion of survey respondents reporting always avoiding contact with other people, stratified by self-reported trust in the CDC (high, some, not much, and none — corresponding to the survey response options “a lot”, “some”, “not too much”, and “not at all”). The four right-hand panels show the same outcome stratified by age, gender, income, and educational attainment. Points represent survey-weighted estimates and vertical bars denote 95% confidence intervals, based on *20* waves of the CHIP50 Project survey. Dashed vertical lines mark key pandemic milestones with potential impact on protective behaviors. *Death toll exceeds 100k* represents the date the officially recorded COVID-19 death toll in the US exceeded 100,000 deaths (May 27, 2020). *CDC universal mask guidance* (July 2020) marks the US CDC’s recommendation that all individuals use face masks in indoor public settings. *Vaccine availability* (spring 2021) denotes the point at which COVID-19 vaccines became widely available to the US population without the eligibility restrictions imposed by the earlier prioritization strategies, reached for all adults in approximately April 2021. *Vaccination coverage 70%* (summer 2021) marks the point at which at least 70% of US adults had received one or more doses of a COVID-19 vaccine, reached in early August 2021. *Omicron dominant* (December 2021) marks the appearance and rapid rise to dominance of the Omicron (B.1.1.529) variant among US infections. *At-home tests widely available* (January 2022) marks the point at which rapid antigen self-tests became broadly accessible to the US public, coinciding with the launch of the federal free-distribution program and the requirement that insurers cover at-home tests.

**Fig S6 | Temporal trajectory of avoiding contacts, stratified by trust in the CDC and by secondary sociodemographic predictors (Political Affiliation, Race/Ethnicity, Employment status, Urbanicity).**

The left panel shows the temporal evolution of the proportion of survey respondents reporting always avoiding contact with other people, stratified by self-reported trust in the CDC (high, some, not much, and none — corresponding to the survey response options “a lot”, “some”, “not too much”, and “not at all”). The four right-hand panels show the same outcome stratified by political affiliation, race/ethnicity, employment status, and urbanicity. Points represent survey-weighted estimates and vertical bars denote 95% confidence intervals, based on *20* waves of the CHIP50 Project survey. Dashed vertical lines mark key pandemic milestones with potential impact on protective behaviors. *Death toll exceeds 100k* represents the date the officially recorded COVID-19 death toll in the US exceeded 100,000 deaths (May 27, 2020). *CDC universal mask guidance* (July 2020) marks the US CDC’s recommendation that all individuals use face masks in indoor public settings. *Vaccine availability* (spring 2021) denotes the point at which COVID-19 vaccines became widely available to the US population without the eligibility restrictions imposed by the earlier prioritization strategies, reached for all adults in approximately April 2021. *Vaccination coverage 70%* (summer 2021) marks the point at which at least 70% of US adults had received one or more doses of a COVID-19 vaccine, reached in early August 2021. *Omicron dominant* (December 2021) marks the appearance and rapid rise to dominance of the Omicron (B.1.1.529) variant among US infections. *At-home tests widely available* (January 2022) marks the point at which rapid antigen self-tests became broadly accessible to the US public, coinciding with the launch of the federal free-distribution program and the requirement that insurers cover at-home tests.

**Fig S7 | Temporal trajectory of avoiding crowds, stratified by trust in the CDC and by core sociodemographic predictors (Age, Gender, Income, Education).**

The left panel shows the temporal evolution of the proportion of survey respondents reporting always avoiding public or crowded places, stratified by self-reported trust in the CDC (high, some, not much, and none — corresponding to the survey response options “a lot”, “some”, “not too much”, and “not at all”). The four right-hand panels show the same outcome stratified by age, gender, income, and educational attainment. Points represent survey-weighted estimates and vertical bars denote 95% confidence intervals, based on *20* waves of the CHIP50 Project survey. Dashed vertical lines mark key pandemic milestones with potential impact on protective behaviors. *Death toll exceeds 100k* represents the date the officially recorded COVID-19 death toll in the US exceeded 100,000 deaths (May 27, 2020). *CDC universal mask guidance* (July 2020) marks the US CDC’s recommendation that all individuals use face masks in indoor public settings. *Vaccine availability* (spring 2021) denotes the point at which COVID-19 vaccines became widely available to the US population without the eligibility restrictions imposed by the earlier prioritization strategies, reached for all adults in approximately April 2021. *Vaccination coverage 70%* (summer 2021) marks the point at which at least 70% of US adults had received one or more doses of a COVID-19 vaccine, reached in early August 2021. *Omicron dominant* (December 2021) marks the appearance and rapid rise to dominance of the Omicron (B.1.1.529) variant among US infections. *At-home tests widely available* (January 2022) marks the point at which rapid antigen self-tests became broadly accessible to the US public, coinciding with the launch of the federal free-distribution program and the requirement that insurers cover at-home tests.

**Fig S8 | Temporal trajectory of avoiding crowds, stratified by trust in the CDC and by secondary sociodemographic predictors (Political Affiliation, Race/Ethnicity, Employment status, Urbanicity).**

The left panel shows the temporal evolution of the proportion of survey respondents reporting always avoiding public or crowded places, stratified by self-reported trust in the CDC (high, some, not much, and none — corresponding to the survey response options “a lot”, “some”, “not too much”, and “not at all”). The four right-hand panels show the same outcome stratified by political affiliation, race/ethnicity, employment status, and urbanicity. Points represent survey-weighted estimates and vertical bars denote 95% confidence intervals, based on *20* waves of the CHIP50 Project survey. Dashed vertical lines mark key pandemic milestones with potential impact on protective behaviors. *Death toll exceeds 100k* represents the date the officially recorded COVID-19 death toll in the US exceeded 100,000 deaths (May 27, 2020). *CDC universal mask guidance* (July 2020) marks the US CDC’s recommendation that all individuals use face masks in indoor public settings. *Vaccine availability* (spring 2021) denotes the point at which COVID-19 vaccines became widely available to the US population without the eligibility restrictions imposed by the earlier prioritization strategies, reached for all adults in approximately April 2021. *Vaccination coverage 70%* (summer 2021) marks the point at which at least 70% of US adults had received one or more doses of a COVID-19 vaccine, reached in early August 2021. *Omicron dominant* (December 2021) marks the appearance and rapid rise to dominance of the Omicron (B.1.1.529) variant among US infections. *At-home tests widely available* (January 2022) marks the point at which rapid antigen self-tests became broadly accessible to the US public, coinciding with the launch of the federal free-distribution program and the requirement that insurers cover at-home tests.

**Fig S9 | Temporal trajectory of vaccination with at least one dose of the COVID-19 vaccine, stratified by trust in the CDC and by core sociodemographic predictors (Age, Gender, Income, Education).**

The left panel shows the temporal evolution of the proportion of survey respondents reporting to have received a COVID-19 vaccine, stratified by self-reported trust in the CDC (high, some, not much, and none — corresponding to the survey response options “a lot”, “some”, “not too much”, and “not at all”). The four right-hand panels show the same outcome stratified by age, gender, income, and educational attainment. Points represent survey-weighted estimates and vertical bars denote 95% confidence intervals, based on *20* waves of the CHIP50 Project survey. Dashed vertical lines mark key pandemic milestones with potential impact on protective behaviors. *Death toll exceeds 100k* represents the date the officially recorded COVID-19 death toll in the US exceeded 100,000 deaths (May 27, 2020). *CDC universal mask guidance* (July 2020) marks the US CDC’s recommendation that all individuals use face masks in indoor public settings. *Vaccine availability* (spring 2021) denotes the point at which COVID-19 vaccines became widely available to the US population without the eligibility restrictions imposed by the earlier prioritization strategies, reached for all adults in approximately April 2021. *Vaccination coverage 70%* (summer 2021) marks the point at which at least 70% of US adults had received one or more doses of a COVID-19 vaccine, reached in early August 2021. *Omicron dominant* (December 2021) marks the appearance and rapid rise to dominance of the Omicron (B.1.1.529) variant among US infections. *At-home tests widely available* (January 2022) marks the point at which rapid antigen self-tests became broadly accessible to the US public, coinciding with the launch of the federal free-distribution program and the requirement that insurers cover at-home tests.

**Fig S10 | Temporal trajectory of vaccination with at least one dose of the COVID-19 vaccine, stratified by trust in the CDC and by secondary sociodemographic predictors (Political Affiliation, Race/Ethnicity, Employment status, Urbanicity).**

The left panel shows the temporal evolution of the proportion of survey respondents reporting to have received a COVID-19 vaccine, stratified by self-reported trust in the CDC (high, some, not much, and none — corresponding to the survey response options “a lot”, “some”, “not too much”, and “not at all”). The four right-hand panels show the same outcome stratified by political affiliation, race/ethnicity, employment status, and urbanicity. Points represent survey-weighted estimates and vertical bars denote 95% confidence intervals, based on *20* waves of the CHIP50 Project survey. Dashed vertical lines mark key pandemic milestones with potential impact on protective behaviors. *Death toll exceeds 100k* represents the date the officially recorded COVID-19 death toll in the US exceeded 100,000 deaths (May 27, 2020). *CDC universal mask guidance* (July 2020) marks the US CDC’s recommendation that all individuals use face masks in indoor public settings. *Vaccine availability* (spring 2021) denotes the point at which COVID-19 vaccines became widely available to the US population without the eligibility restrictions imposed by the earlier prioritization strategies, reached for all adults in approximately April 2021. *Vaccination coverage 70%* (summer 2021) marks the point at which at least 70% of US adults had received one or more doses of a COVID-19 vaccine, reached in early August 2021. *Omicron dominant* (December 2021) marks the appearance and rapid rise to dominance of the Omicron (B.1.1.529) variant among US infections. *At-home tests widely available* (January 2022) marks the point at which rapid antigen self-tests became broadly accessible to the US public, coinciding with the launch of the federal free-distribution program and the requirement that insurers cover at-home tests.

**Fig S11 | Pooled pandemic-era adjusted odds ratios for avoiding contact and avoiding crowds, by trust in the CDC and sociodemographic covariates.**

Each column presents adjusted odds ratios (ORs) from a survey-weighted multivariable logistic regression, estimated separately within three pandemic eras — pre-vaccine (waves 1–13; April 2020 – November 2020), vaccine rollout (waves 14–20, December 2020 – December 2021), and Omicron (waves 21–24, December 2021 – September 2022) — for two outcomes: avoiding contact with others (left three columns) and avoiding crowds (right three columns). Point estimates are adjusted ORs and horizontal lines denote 95% confidence intervals, plotted on a log scale; the dashed vertical line marks the null (OR = 1). Rows are grouped by trust and covariates, color-coded by social-determinant-of-health (SDH) group, and ordered hierarchically by the magnitude of association (|log OR|) so that the most strongly associated predictors appear at the top. The primary exposure, trust in the CDC (gold), contrasts high, some, and not-much trust against no trust (reference). Two model specifications are displayed: Specification A (dots) pools survey waves according to pandemic era, whereas Specification B (triangles) adds wave-fixed effects to absorb within-era temporal variation (the wave-indicator coefficients are not shown for conciseness). Estimates are based on 20 waves of the CHIP50 Project survey (waves 1–24, excluding waves 4, 6, 8, and 15).

**Fig S12 | Pooled pandemic-era adjusted odds ratios for mask wearing and handwashing, by trust in the CDC and sociodemographic covariates.** Each column presents adjusted odds ratios (ORs) from a survey-weighted multivariable logistic regression, estimated separately within three pandemic eras — pre-vaccine (waves 1–13; April 2020 – November 2020), vaccine rollout (waves 14–20, December 2020 – December 2021), and Omicron (waves 21–24, December 2021 – September 2022) — for two outcomes: mask wearing (left three columns) and handwashing (right three columns). Point estimates are adjusted ORs and horizontal lines denote 95% confidence intervals, plotted on a log scale; the dashed vertical line marks the null (OR = 1). Rows are grouped by trust and covariates, color-coded by social-determinant-of-health (SDH) group, and ordered hierarchically by the magnitude of association (|log OR|) so that the most strongly associated predictors appear at the top. The primary exposure, trust in the CDC (gold), contrasts high, some, and not-much trust against no trust (reference). Two model specifications are displayed: Specification A (dots) pools survey waves according to pandemic era, whereas Specification B (triangles) adds wave-fixed effects to absorb within-era temporal variation (the wave-indicator coefficients are not shown for conciseness). Estimates are based on 20 waves of the CHIP50 Project survey (waves 1–24, excluding waves 4, 6, 8, and 15).

**Fig S13 | Pooled pandemic-era adjusted odds ratios for vaccination (1+ dose(s)) and visiting a doctor or hospital, by trust in the CDC and sociodemographic covariates.**

Each column presents adjusted odds ratios (ORs) from a survey-weighted multivariable logistic regression, estimated separately within three pandemic eras — pre-vaccine (waves 1–13; April 2020 – November 2020), vaccine rollout (waves 14–20, December 2020 – December 2021), and Omicron (waves 21–24, December 2021 – September 2022) — for two outcomes: getting at least one dose of a COVID-19 vaccine (left three columns) and visiting a doctor or hospital within the last 24 hours (right three columns). Point estimates are adjusted ORs and horizontal lines denote 95% confidence intervals, plotted on a log scale; the dashed vertical line marks the null (OR = 1). Rows are grouped by trust and covariates, color-coded by social-determinant-of-health (SDH) group, and ordered hierarchically by the magnitude of association (|log OR|) so that the most strongly associated predictors appear at the top. The primary exposure, trust in the CDC (gold), contrasts high, some, and not-much trust against no trust (reference). Two model specifications are displayed: Specification A (dots) pools survey waves according to pandemic era, whereas Specification B (triangles) adds wave-fixed effects to absorb within-era temporal variation (the wave-indicator coefficients are not shown for conciseness). Estimates are based on 20 waves of the CHIP50 Project survey (waves 1–24, excluding waves 4, 6, 8, and 15).

**Fig S14 | Pooled pandemic-era adjusted odds ratios for avoiding contact and avoiding crowds, by trust in banks and sociodemographic covariates.**

Each column presents adjusted odds ratios (ORs) from a survey-weighted multivariable logistic regression, estimated separately within three pandemic eras — pre-vaccine (waves 1–13; April 2020 – November 2020), vaccine rollout (waves 14–20, December 2020 – December 2021), and Omicron (waves 21–24, December 2021 – September 2022) — for two outcomes: avoiding contact with others (left three columns) and avoiding crowds (right three columns). Point estimates are adjusted ORs and horizontal lines denote 95% confidence intervals, plotted on a log scale; the dashed vertical line marks the null (OR = 1). Rows are grouped by trust and covariates, color-coded by social-determinant-of-health (SDH) group, and ordered hierarchically by the magnitude of association (|log OR|) so that the most strongly associated predictors appear at the top. The negative control exposure, trust in banks (gold), and the primary exposure, trust in the CDC (dark gray), contrast high, some, and not-much trust against no trust (reference). Two model specifications are displayed: Specification A (dots) pools survey waves according to pandemic era, whereas Specification B (triangles) adds wave-fixed effects to absorb within-era temporal variation (the wave-indicator coefficients are not shown for conciseness). Estimates are based on 20 waves of the CHIP50 Project survey (waves 1–24, excluding waves 4, 6, 8, and 15).

**Fig S15 | Pooled pandemic-era adjusted odds ratios for mask wearing and handwashing, by trust in banks and sociodemographic covariates.** Each column presents adjusted odds ratios (ORs) from a survey-weighted multivariable logistic regression, estimated separately within three pandemic eras — pre-vaccine (waves 1–13; April 2020 – November 2020), vaccine rollout (waves 14–20, December 2020 – December 2021), and Omicron (waves 21–24, December 2021 – September 2022) — for two outcomes: mask wearing (left three columns) and handwashing (right three columns). Point estimates are adjusted ORs and horizontal lines denote 95% confidence intervals, plotted on a log scale; the dashed vertical line marks the null (OR = 1). Rows are grouped by trust and covariates, color-coded by social-determinant-of-health (SDH) group, and ordered hierarchically by the magnitude of association (|log OR|) so that the most strongly associated predictors appear at the top. The negative control exposure, trust in banks (gold), and the primary exposure, trust in the CDC (dark gray), contrast high, some, and not-much trust against no trust (reference). Two model specifications are displayed: Specification A (dots) pools survey waves according to pandemic era, whereas Specification B (triangles) adds wave-fixed effects to absorb within-era temporal variation (the wave-indicator coefficients are not shown for conciseness). Estimates are based on 20 waves of the CHIP50 Project survey (waves 1–24, excluding waves 4, 6, 8, and 15).

**Fig S16 | Pooled pandemic-era adjusted odds ratios for vaccination (1+ dose(s)) and visiting a doctor or hospital, by trust in banks and sociodemographic covariates.**

Each column presents adjusted odds ratios (ORs) from a survey-weighted multivariable logistic regression, estimated separately within three pandemic eras — pre-vaccine (waves 1–13; April 2020 – November 2020), vaccine rollout (waves 14–20, December 2020 – December 2021), and Omicron (waves 21–24, December 2021 – September 2022) — for two outcomes: getting at least one dose of a COVID-19 vaccine (left three columns) and visiting a doctor or hospital within the last 24 hours (right three columns). Point estimates are adjusted ORs and horizontal lines denote 95% confidence intervals, plotted on a log scale; the dashed vertical line marks the null (OR = 1). Rows are grouped by trust and covariates, color-coded by social-determinant-of-health (SDH) group, and ordered hierarchically by the magnitude of association (|log OR|) so that the most strongly associated predictors appear at the top. The negative control exposure, trust in banks (gold), and the primary exposure, trust in the CDC (dark gray), contrast high, some, and not-much trust against no trust (reference). Two model specifications are displayed: Specification A (dots) pools survey waves according to pandemic era, whereas Specification B (triangles) adds wave-fixed effects to absorb within-era temporal variation (the wave-indicator coefficients are not shown for conciseness). Estimates are based on 20 waves of the CHIP50 Project survey (waves 1–24, excluding waves 4, 6, 8, and 15).

**Fig S17 | Adjusted high- versus no-trust risk differences for trust in the CDC across alternative model specifications.**

Each panel shows the temporal trajectory of the adjusted risk difference for a given protective or health-seeking behavior between respondents reporting high trust vs. no trust in the CDC, estimated from survey-weighted multivariable logistic regressions fit separately within each CHIP50 wave. The adjusted risk difference is expressed in percentage points – positive values indicate higher behavioral prevalence among high-trust respondents. The six panels correspond to the four primary protective behaviors (always wearing a mask, always avoiding contact, always avoiding crowds, and frequent handwashing) and two additional outcomes (receipt of at least one vaccine dose and visiting a doctor or hospital). Within each panel, three model specifications are compared: the primary specification, which adjusts for age, gender, race/ethnicity, education, household income, urbanicity (Rural–Urban Commuting Area codes), partisan leaning, ideology, U.S. Census region, and employment; a sensitivity specification with party and ideology removed from the covariate set; and a sensitivity specification additionally conditioning on a continuous index of generalized trust in others. Lines connect the survey-weighted point estimates across waves and shaded bands denote 95% confidence intervals. Estimates are based on the included CHIP50 Project survey waves (waves 1–24, excluding waves 4, 6, 8, and 15 owing to insufficient sample size); vaccination items were fielded from approximately wave 14 onward. The near-identity of the generalized-trust–adjusted trajectory (green) to the primary trajectory (blue) across all outcomes indicates that the CDC trust contrast likely reflects trust specific to public-health institutions beyond the influence of a dispositional propensity toward interpersonal trust. Note that the y-axis scale differs across panels.

**Fig S18 | Concordance of adjusted trust-in-CDC risk differences with and without adjustment for general trust**.

Each point represents one behavioral outcome at one survey wave (110 outcome × wave estimates across CHIP50 Project waves 1–24, excluding waves 4, 6, 8, and 15). The horizontal axis (x-axis) represents the adjusted high- versus no-trust difference by trust in the CDC estimated from the primary multivariable model (adjusting for age, gender, race/ethnicity, education, household income, urbanicity, partisan leaning, ideology, U.S. Census region, and employment), and the vertical axis (y-axis) represents the corresponding estimate from the sensitivity model that additionally conditions on a continuous index of generalized trust in others; both risk differences are expressed in percentage points. Points are colored by behavioral outcome (always wearing a mask, always avoiding contact, always avoiding crowds, frequent handwashing, receipt of at least one vaccine dose, and visiting a doctor or hospital). The dashed diagonal line is the line of identity (y = x); points falling on this line are unchanged by the additional adjustment for generalized trust. Estimates cluster tightly along the line of identity across the full range of effect sizes — from near-zero differences for doctor or hospital visits to differences exceeding 40 percentage points for vaccination — with a Pearson correlation of 0.99 between the two specifications, a median absolute change of 0.8 percentage points (maximum 3.6 percentage points), and a median relative change of −2% (interquartile range −6.6% to +1.8%). This near-identity indicates that the estimated association between trust in the CDC and protective behavior is essentially invariant to adjustment for a dispositional propensity toward interpersonal trust, reflecting trust specifically in public-health institutions rather than generalized social trust.

**Fig S19 | Temporal trajectories of institutional trust during the COVID-19 pandemic.**

Each panel shows the temporal evolution of the survey-weighted proportion of respondents reporting each of four levels of trust — high, some, not much, and none, corresponding to the survey response options “a lot”, “some”, “not too much”, and “not at all” — in a given institution: the US Centers for Disease Control and Prevention (CDC), doctors and hospitals, state governments, and banks. Banks are included as a non-health institution serving as the negative-control exposure in the adjusted analyses (see Methods). Within each panel, lines connect the survey-weighted point estimates for a given trust level across waves and shaded bands denote 95% confidence intervals, based on 20 waves of the CHIP50 Project survey spanning April 2020 to August 2022 (waves 1–24, excluding waves 4, 6, 8, and 15 owing to insufficient sample size for the analyzed variables). Across all four institutions, the relative ordering and separation of the trust strata remained broadly stable over the course of the pandemic, indicating that the relative composition of high- versus low-trust groups was approximately temporally invariant. Dashed vertical lines mark key pandemic milestones: *Death toll exceeds 100k* represents the date the officially recorded COVID-19 death toll in the US exceeded 100,000 deaths (May 27, 2020). *CDC universal mask guidance* (July 2020) marks the US CDC’s recommendation that all individuals use face masks in indoor public settings. *Vaccine availability* (spring 2021) denotes the point at which COVID-19 vaccines became widely available to the US population without the eligibility restrictions imposed by earlier prioritization strategies. *Vaccination coverage 70%* (summer 2021) marks the point at which at least 70% of US adults had received one or more doses of a COVID-19 vaccine, reached in early August 2021. *Omicron dominant* (January 2022) marks the rise of the Omicron (B.1.1.529) variant to dominance among US infections. *At-home test kits available* (January 2022) marks the point at which rapid antigen self-tests became broadly accessible to the US public, coinciding with the launch of the federal free-distribution program and the requirement that insurers cover at-home tests.

## Notes

### Author Declarations

The institutional review board of Harvard University deemed this study exempt as only deidentified data were used and no participant contact was required.

