## Supplementary Materials for "Trust as a Hidden Driver of Epidemic Dynamics: A Missing Parameter in Compartmental Disease Transmission Models"

**Supplementary Table S1: Leave-one-block-out analysis of explanatory variance in the fully adjusted model (M4).**

**Supplementary Figures S1-S10: Descriptive Trajectories**

**Supplementary Figures S11-S19: Adjusted Trust Estimates**

**Supplementary Table S1: Leave-one-block-out analysis of explanatory variance in the fully adjusted model (M4).**

| **Block removed** | **ΔAIC penalty (median [IQR])** | **ΔBrier penalty (median [IQR])** |
| --- | --- | --- |
| ***Mask wearing — Pre-vaccine era (waves = 9; median n = 18,991)*** | | |
| Trust in the CDC | +879 [413, 955] | +0.0094 [0.0068, 0.0103] |
| Political identity (party & ideology) | +379 [239, 583] | +0.0040 [0.0035, 0.0062] |
| Demographics (age, race/ethnicity, gender) | +318 [288, 373] | +0.0036 [0.0029, 0.0043] |
| Geographic context (urbanicity & region) | +302 [144, 358] | +0.0035 [0.0014, 0.0047] |
| Socioeconomic status (education, income, employment) | +34 [24, 50] | +0.0006 [0.0004, 0.0009] |
| ***Mask wearing — Vaccine rollout era (waves = 7; median n = 23,527)*** | | |
| Trust in the CDC | +1,315 [1,128, 1,503] | +0.0109 [0.0099, 0.0119] |
| Political identity (party & ideology) | +455 [394, 519] | +0.0042 [0.0035, 0.0043] |
| Demographics (age, race/ethnicity, gender) | +363 [333, 625] | +0.0025 [0.0022, 0.0058] |
| Geographic context (urbanicity & region) | +190 [146, 243] | +0.0018 [0.0012, 0.0023] |
| Socioeconomic status (education, income, employment) | +17 [6, 39] | +0.0002 [0.0002, 0.0005] |
| ***Mask wearing — Omicron era (waves = 4; median n = 23,986)*** | | |
| Demographics (age, race/ethnicity, gender) | +738 [626, 863] | +0.0076 [0.0066, 0.0087] |
| Political identity (party & ideology) | +494 [394, 578] | +0.0037 [0.0029, 0.0049] |
| Trust in the CDC | +434 [243, 719] | +0.0040 [0.0026, 0.0059] |
| Geographic context (urbanicity & region) | +163 [134, 270] | +0.0014 [0.0012, 0.0024] |
| Socioeconomic status (education, income, employment) | +92 [62, 120] | +0.0010 [0.0007, 0.0013] |
| ***Avoiding contact — Pre-vaccine era (waves = 9; median n = 19,045)*** | | |
| Trust in the CDC | +331 [297, 421] | +0.0044 [0.0037, 0.0051] |
| Demographics (age, race/ethnicity, gender) | +200 [161, 292] | +0.0030 [0.0026, 0.0034] |
| Political identity (party & ideology) | +158 [119, 247] | +0.0020 [0.0017, 0.0030] |
| Socioeconomic status (education, income, employment) | +124 [110, 170] | +0.0019 [0.0017, 0.0020] |
| Geographic context (urbanicity & region) | +63 [51, 80] | +0.0009 [0.0006, 0.0011] |
| ***Avoiding contact — Vaccine rollout era (waves = 7; median n = 23,592)*** | | |
| Trust in the CDC | +403 [392, 488] | +0.0036 [0.0031, 0.0045] |
| Political identity (party & ideology) | +363 [188, 409] | +0.0034 [0.0015, 0.0039] |
| Demographics (age, race/ethnicity, gender) | +248 [210, 277] | +0.0023 [0.0021, 0.0027] |
| Socioeconomic status (education, income, employment) | +147 [142, 160] | +0.0016 [0.0014, 0.0018] |
| Geographic context (urbanicity & region) | +43 [36, 59] | +0.0004 [0.0003, 0.0006] |
| ***Avoiding contact — Omicron era (waves = 4; median n = 24,024)*** | | |
| Demographics (age, race/ethnicity, gender) | +259 [192, 331] | +0.0026 [0.0021, 0.0028] |
| Socioeconomic status (education, income, employment) | +217 [187, 262] | +0.0020 [0.0017, 0.0022] |
| Trust in the CDC | +182 [133, 239] | +0.0014 [0.0012, 0.0016] |
| Political identity (party & ideology) | +177 [120, 218] | +0.0009 [0.0007, 0.0013] |
| Geographic context (urbanicity & region) | +32 [24, 49] | +0.0003 [0.0003, 0.0005] |
| ***Avoiding crowds — Pre-vaccine era (waves = 9; median n = 19,019)*** | | |
| Trust in the CDC | +392 [350, 480] | +0.0053 [0.0048, 0.0060] |
| Political identity (party & ideology) | +226 [109, 322] | +0.0027 [0.0019, 0.0040] |
| Demographics (age, race/ethnicity, gender) | +201 [153, 297] | +0.0027 [0.0025, 0.0036] |
| Socioeconomic status (education, income, employment) | +100 [98, 114] | +0.0015 [0.0013, 0.0016] |
| Geographic context (urbanicity & region) | +37 [28, 47] | +0.0006 [0.0004, 0.0007] |
| ***Avoiding crowds — Vaccine rollout era (waves = 7; median n = 23,549)*** | | |
| Trust in the CDC | +608 [557, 647] | +0.0053 [0.0052, 0.0064] |
| Demographics (age, race/ethnicity, gender) | +269 [241, 280] | +0.0025 [0.0023, 0.0030] |
| Political identity (party & ideology) | +248 [196, 360] | +0.0024 [0.0018, 0.0035] |
| Socioeconomic status (education, income, employment) | +131 [108, 139] | +0.0014 [0.0013, 0.0015] |
| Geographic context (urbanicity & region) | +55 [48, 56] | +0.0006 [0.0005, 0.0006] |
| ***Avoiding crowds — Omicron era (waves = 4; median n = 23,982)*** | | |
| Trust in the CDC | +288 [204, 371] | +0.0025 [0.0021, 0.0028] |
| Socioeconomic status (education, income, employment) | +261 [206, 321] | +0.0028 [0.0021, 0.0033] |
| Demographics (age, race/ethnicity, gender) | +216 [145, 288] | +0.0024 [0.0020, 0.0027] |
| Political identity (party & ideology) | +191 [137, 235] | +0.0012 [0.0010, 0.0016] |
| Geographic context (urbanicity & region) | +24 [20, 49] | +0.0003 [0.0002, 0.0006] |
| ***Handwashing — Pre-vaccine era (waves = 9; median n = 19,015)*** | | |
| Demographics (age, race/ethnicity, gender) | +360 [331, 406] | +0.0039 [0.0034, 0.0042] |
| Trust in the CDC | +290 [205, 304] | +0.0030 [0.0028, 0.0031] |
| Political identity (party & ideology) | +18 [6, 49] | +0.0003 [0.0001, 0.0005] |
| Geographic context (urbanicity & region) | +14 [10, 23] | +0.0003 [0.0002, 0.0004] |
| Socioeconomic status (education, income, employment) | +12 [7, 18] | +0.0003 [0.0003, 0.0004] |
| ***Handwashing — Vaccine rollout era (waves = 7; median n = 23,548)*** | | |
| Demographics (age, race/ethnicity, gender) | +498 [339, 505] | +0.0039 [0.0034, 0.0043] |
| Trust in the CDC | +441 [369, 460] | +0.0041 [0.0035, 0.0043] |
| Political identity (party & ideology) | +51 [29, 59] | +0.0005 [0.0003, 0.0006] |
| Geographic context (urbanicity & region) | +20 [15, 31] | +0.0003 [0.0002, 0.0004] |
| Socioeconomic status (education, income, employment) | +18 [12, 32] | +0.0004 [0.0003, 0.0005] |
| ***Handwashing — Omicron era (waves = 4; median n = 23,979)*** | | |
| Demographics (age, race/ethnicity, gender) | +623 [527, 690] | +0.0067 [0.0063, 0.0068] |
| Trust in the CDC | +284 [231, 315] | +0.0026 [0.0025, 0.0030] |
| Socioeconomic status (education, income, employment) | +37 [23, 52] | +0.0007 [0.0006, 0.0007] |
| Geographic context (urbanicity & region) | +18 [15, 21] | +0.0003 [0.0003, 0.0003] |
| Political identity (party & ideology) | +7 [4, 13] | +0.0002 [0.0001, 0.0002] |

*Each entry summarizes the change in model fit when a single covariate block is removed from the fully adjusted model (M4) and the model is refit on the identical complete-case sample; M4 contains trust in the CDC together with the demographic, socioeconomic, political, and geographic blocks. Values are the median and interquartile range (IQR) of the wave-specific estimates within each pandemic era. The ΔAIC penalty is AIC(reduced model, block removed) − AIC(full model M4); the ΔBrier penalty is Brier(reduced) − Brier(full). Positive values denote a penalty: removing the block degrades model fit (higher AIC) and probabilistic calibration (higher Brier score), so larger positive values indicate greater degradation and therefore more unique information contributed by that block. Within each outcome × era group, blocks are ordered from largest to smallest ΔAIC penalty. Covariate blocks: Trust in the CDC; Demographics (age, race/ethnicity, gender); Socioeconomic status (educational attainment, household income, employment); Political identity (3-level party identification and ideology); Geographic context (RUCA-based urbanicity and U.S. Census region). In design-based likelihood-ratio and Wald tests, removal of each block significantly worsened fit (FDR-adjusted q < 0.05) in all or nearly all waves within each era. Pandemic eras: pre-vaccine, vaccine rollout, and Omicron. AIC, Akaike information criterion; FDR, false discovery rate; IQR, interquartile range; M4, fully adjusted model; RUCA, Rural–Urban Commuting Area.*

**Supplementary figures**

**Fig S1 | Temporal trajectory of mask wearing, stratified by trust in the CDC and by secondary sociodemographic predictors (Political Affiliation, Race/Ethnicity, Employment status, Urbanicity)**

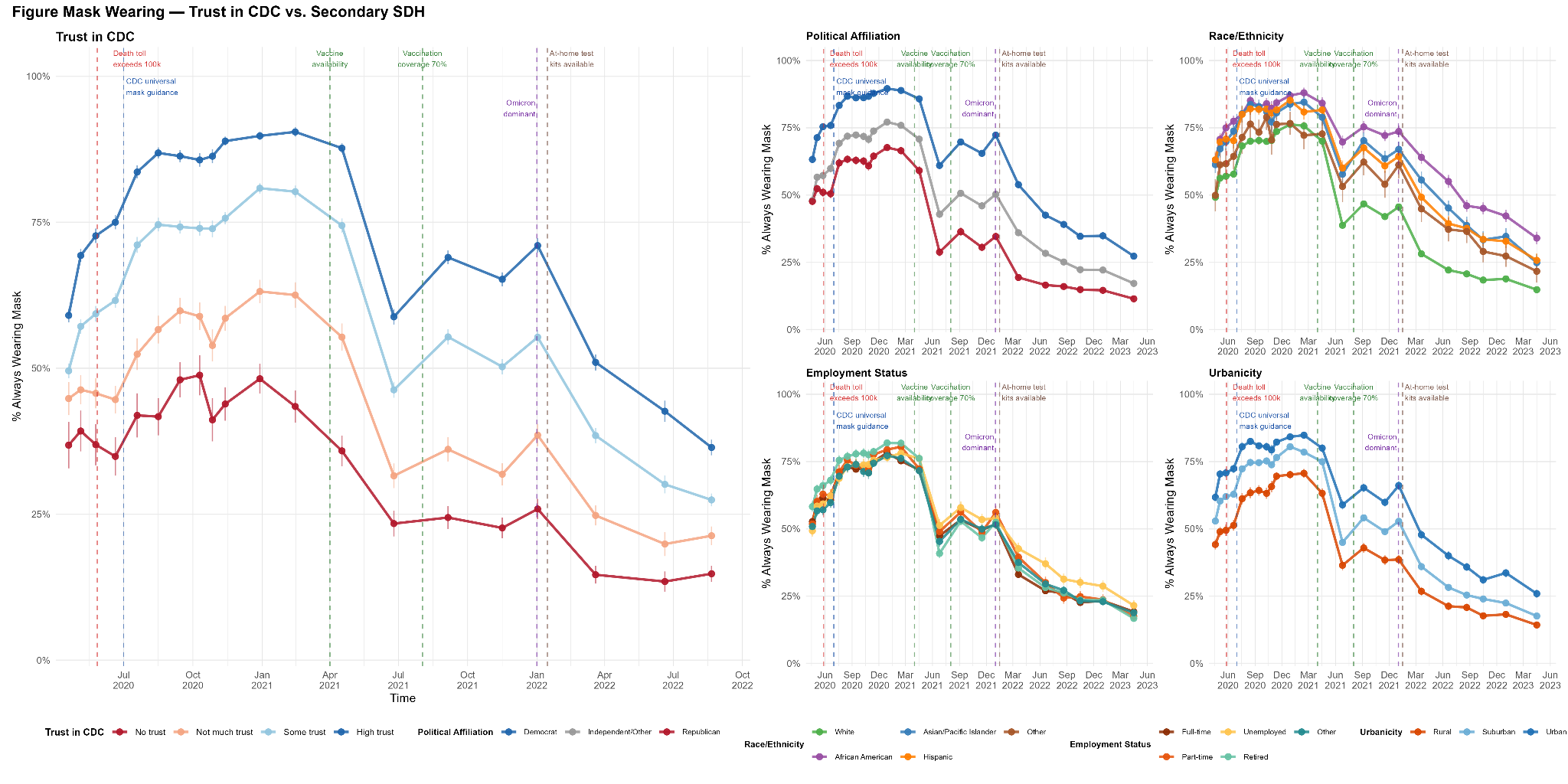

**Fig S2 | Temporal trajectory of handwashing, stratified by trust in the CDC and by secondary sociodemographic predictors (Political Affiliation, Race/Ethnicity, Employment status, Urbanicity).**

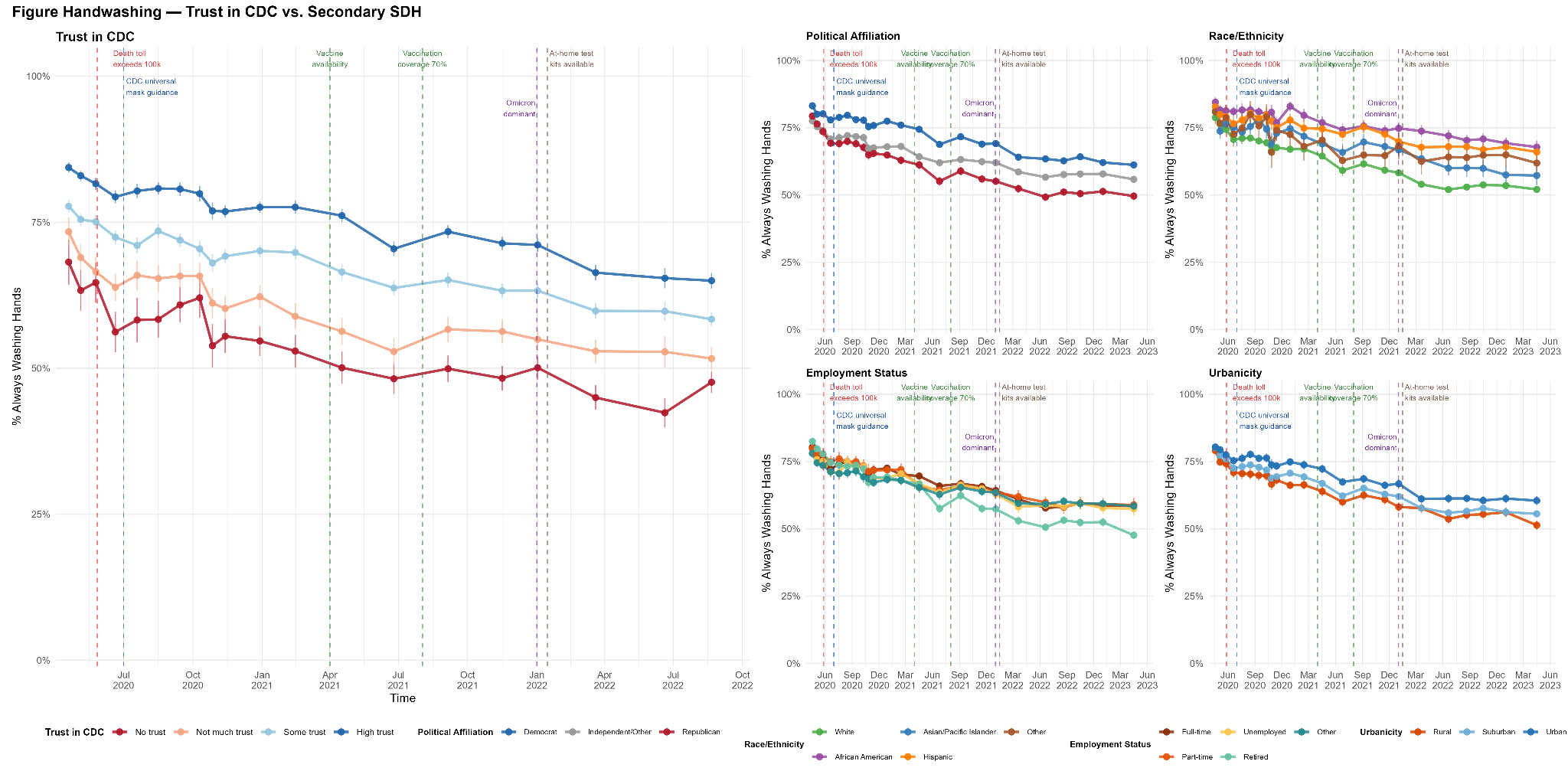

Fig **S3 | Temporal trajectory of mask wearing, stratified by trust in banks (negative control exposure) and by core sociodemographic predictors (Age, Gender, Income, Education).**

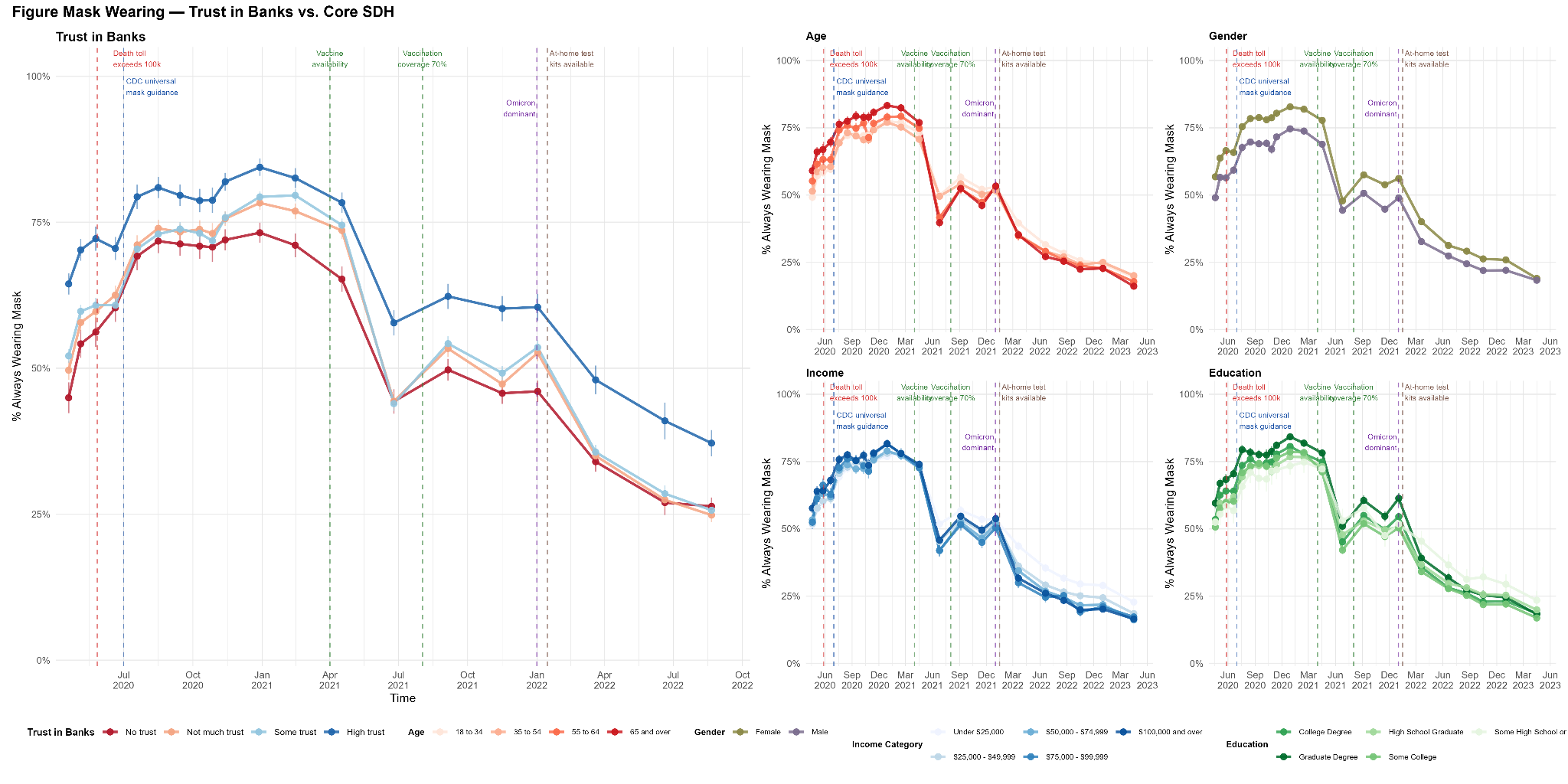

**Fig S4 | Temporal trajectory of mask wearing, stratified by trust in banks (negative control exposure) and by secondary sociodemographic predictors (Political Affiliation, Race/Ethnicity, Employment status, Urbanicity).**

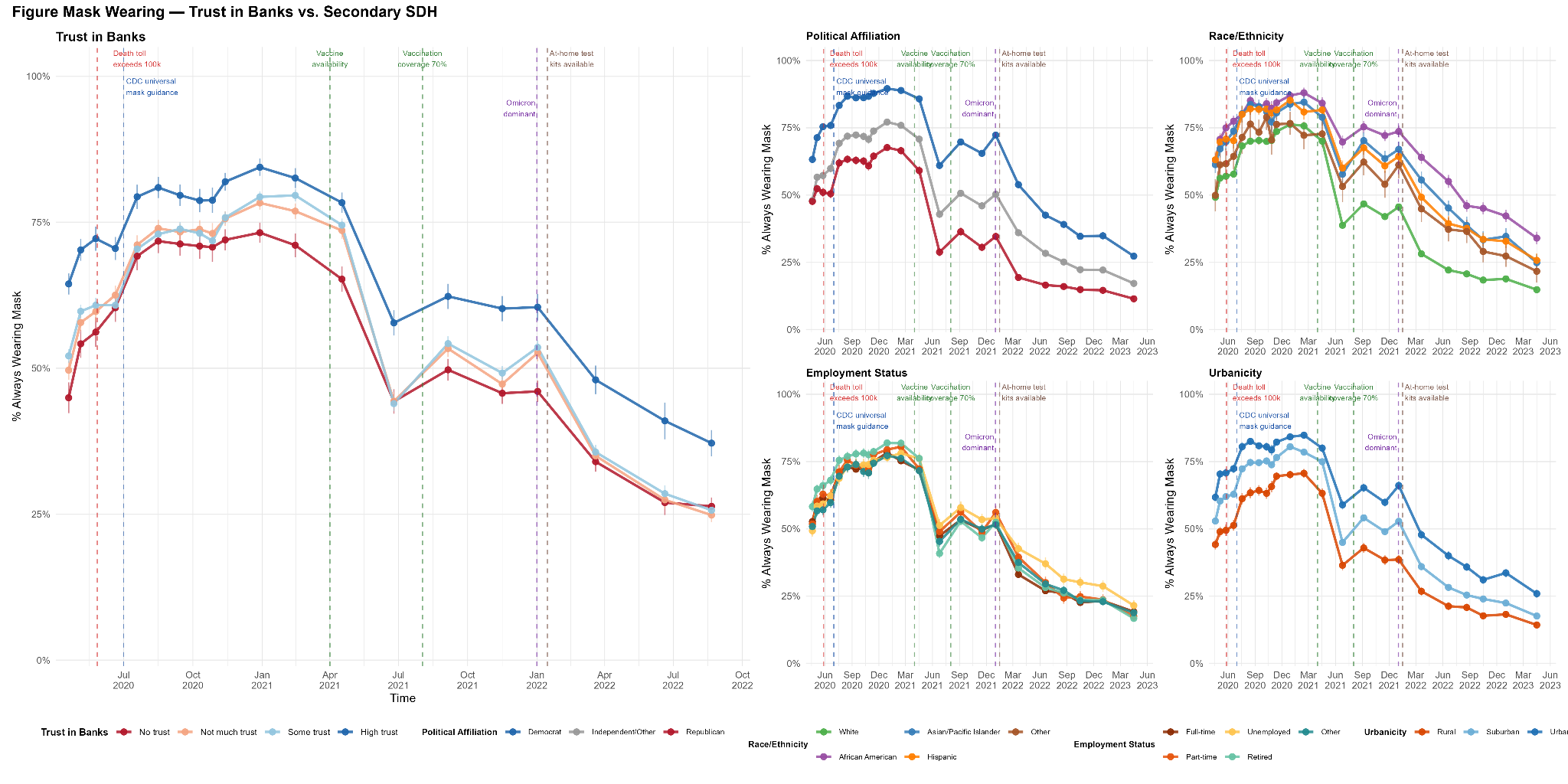

**Fig S5 | Temporal trajectory of avoiding contacts, stratified by trust in the CDC and by core sociodemographic predictors (Age, Gender, Income, Education).**

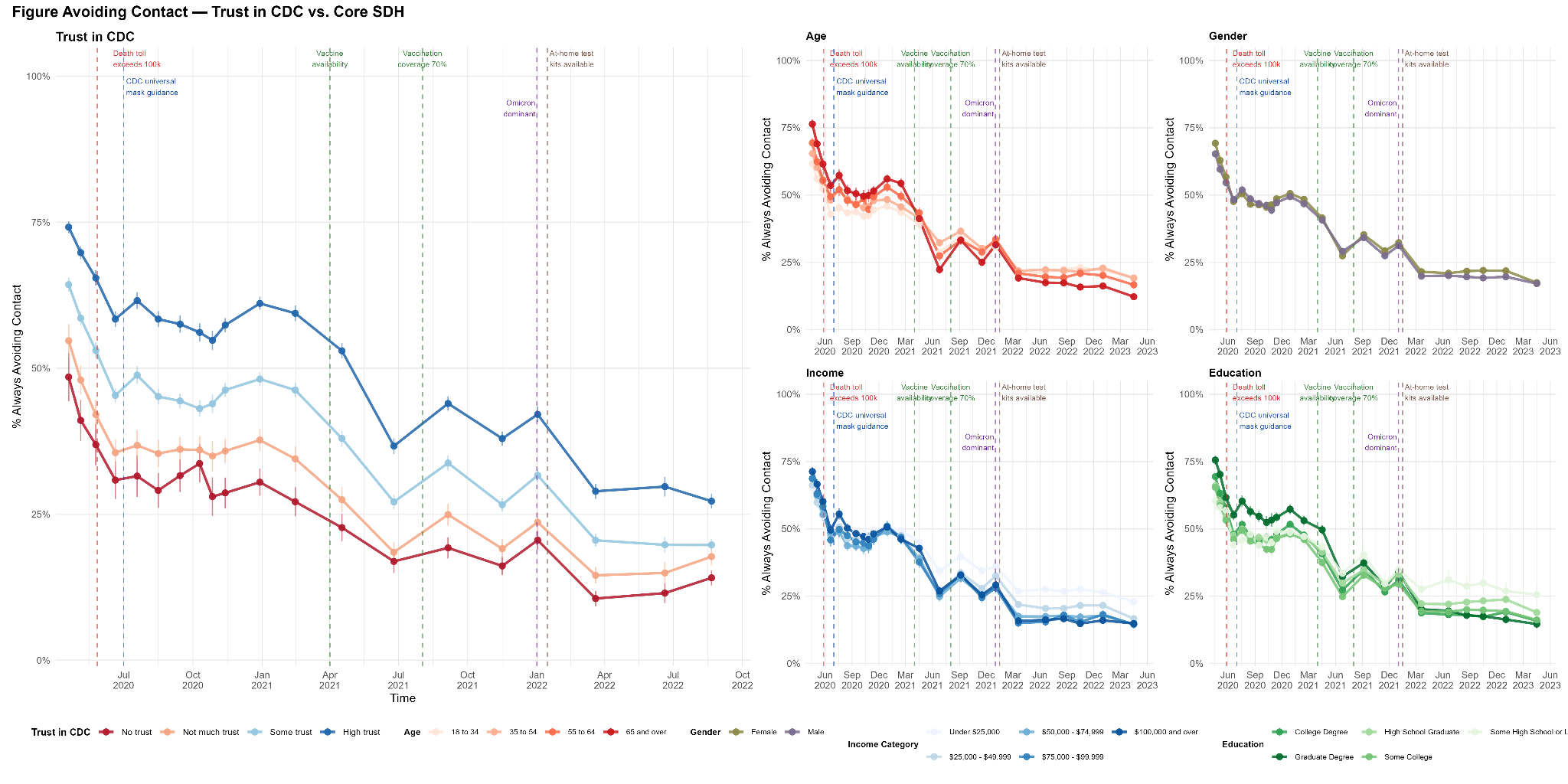

**Fig S6 | Temporal trajectory of avoiding contacts, stratified by trust in the CDC and by secondary sociodemographic predictors (Political Affiliation, Race/Ethnicity, Employment status, Urbanicity).**

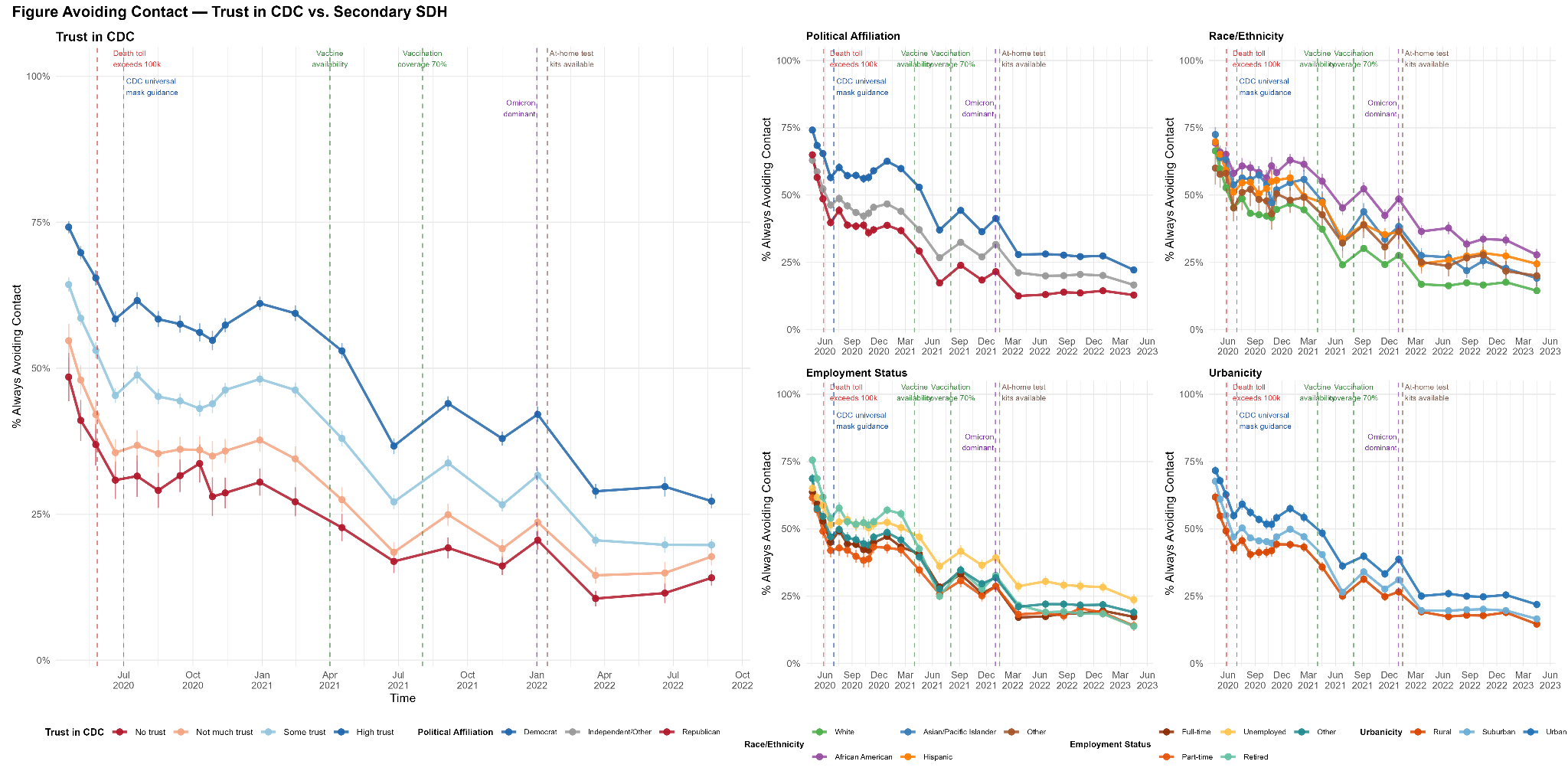

**Fig S7 | Temporal trajectory of avoiding crowds, stratified by trust in the CDC and by core sociodemographic predictors (Age, Gender, Income, Education).**

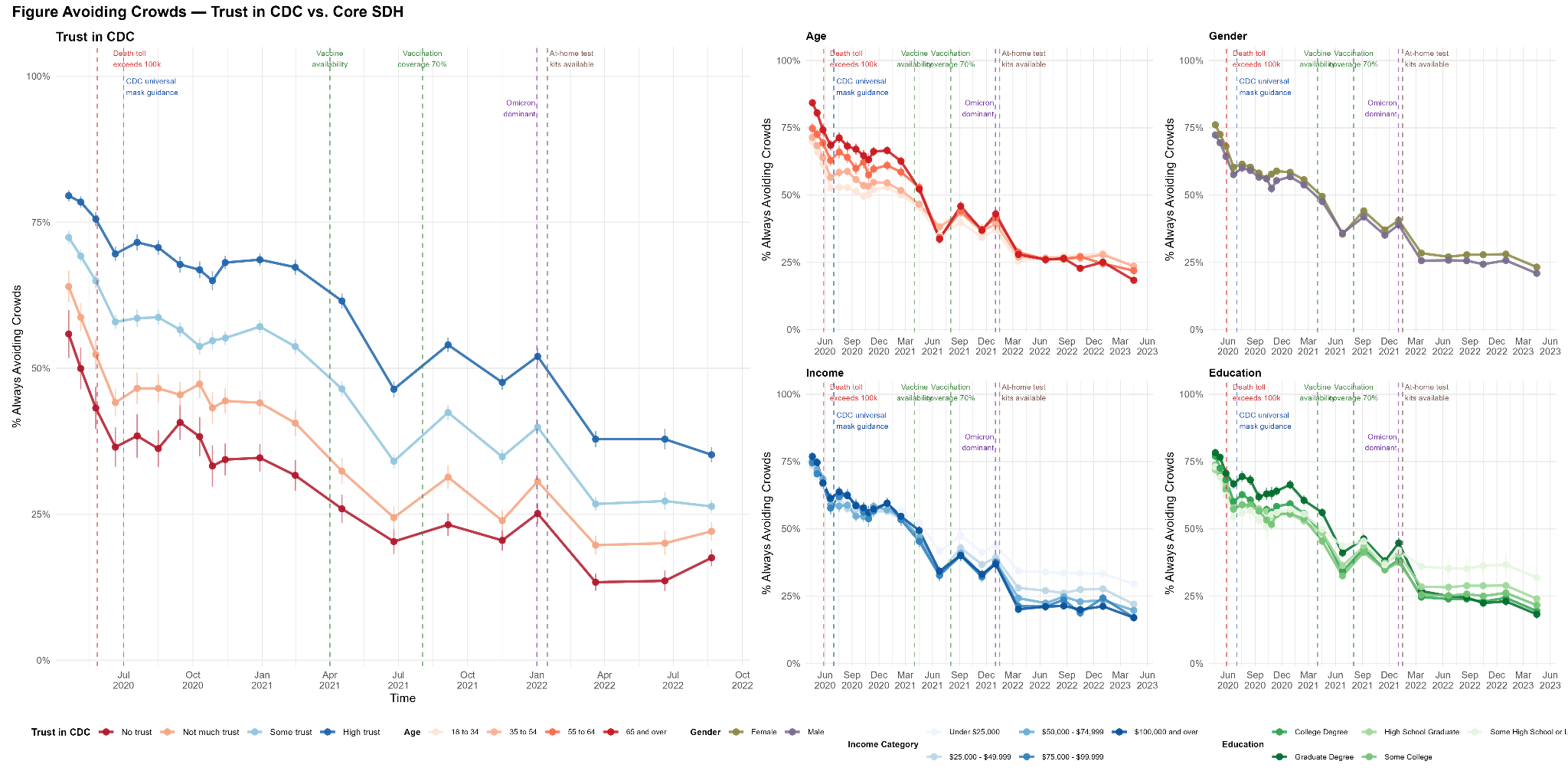

**Fig S8 | Temporal trajectory of avoiding crowds, stratified by trust in the CDC and by secondary sociodemographic predictors (Political Affiliation, Race/Ethnicity, Employment status, Urbanicity).**

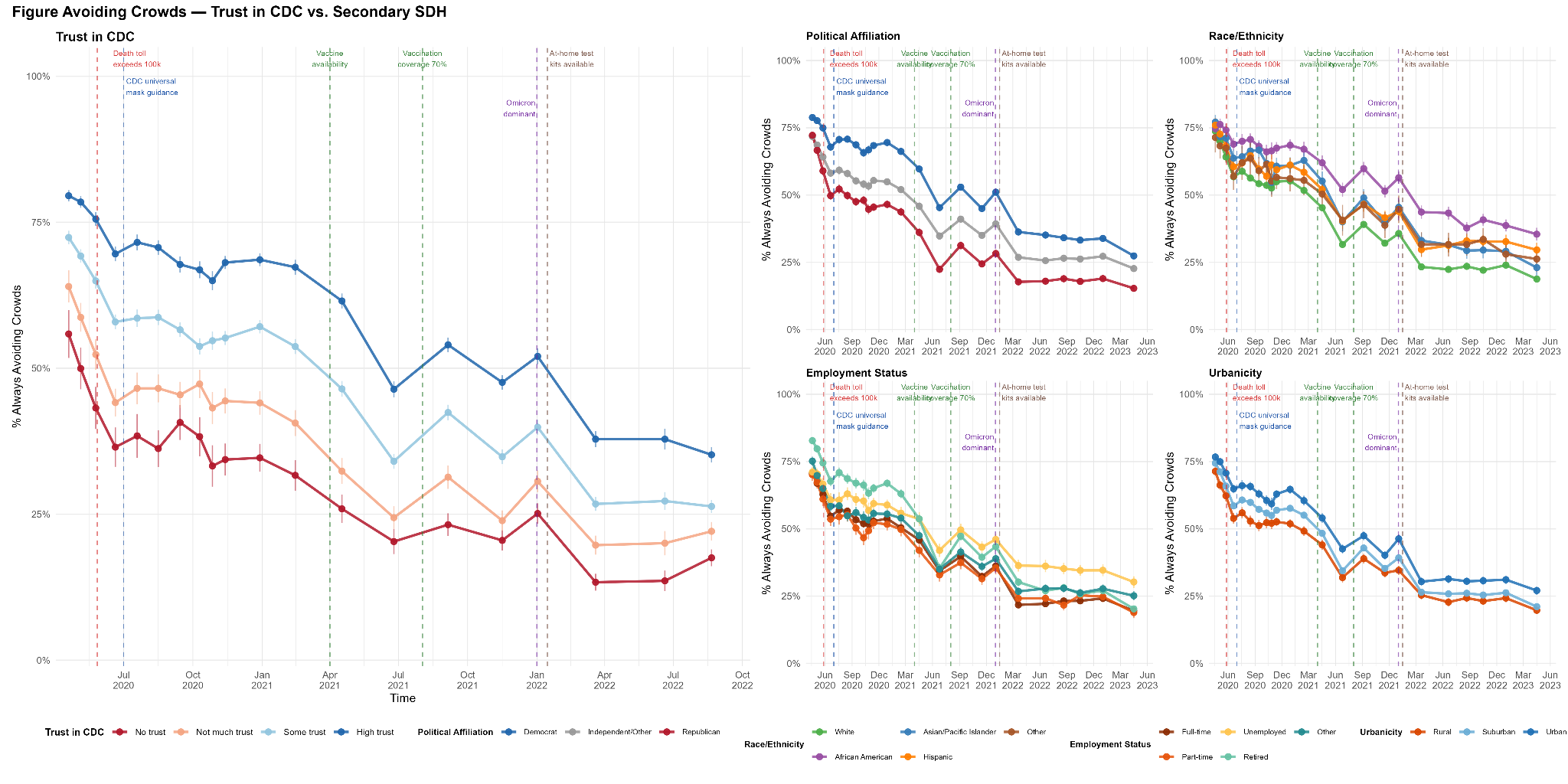

**Fig S9 | Temporal trajectory of vaccination with at least one dose of the COVID-19 vaccine, stratified by trust in the CDC and by core sociodemographic predictors (Age, Gender, Income, Education).**

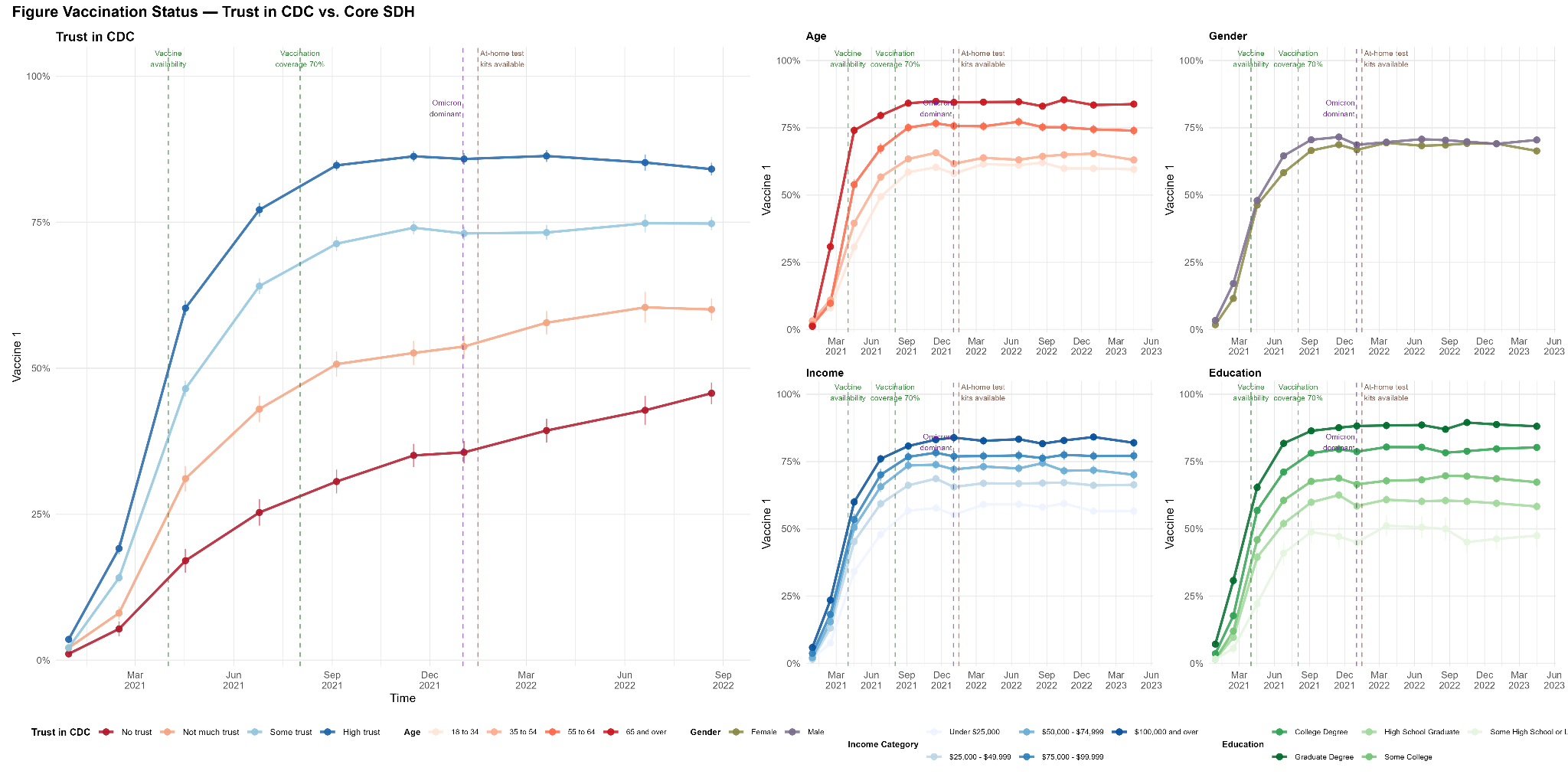

**Fig S10 | Temporal trajectory of vaccination with at least one dose of the COVID-19 vaccine, stratified by trust in the CDC and by secondary sociodemographic predictors (Political Affiliation, Race/Ethnicity, Employment status, Urbanicity).**

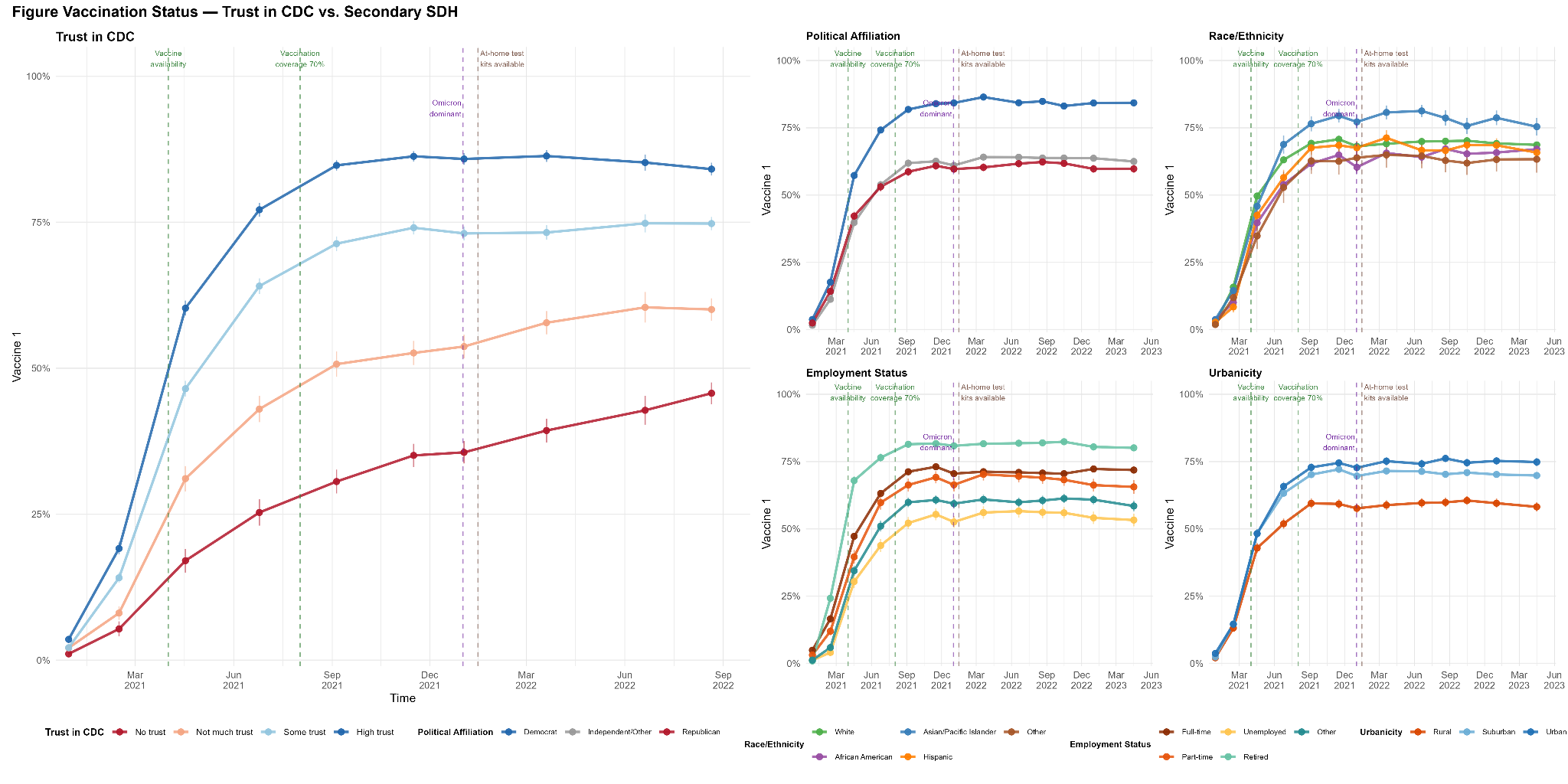

**Fig S11 | Pooled pandemic-era adjusted odds ratios for avoiding contact and avoiding crowds, by trust in the CDC and sociodemographic covariates.**

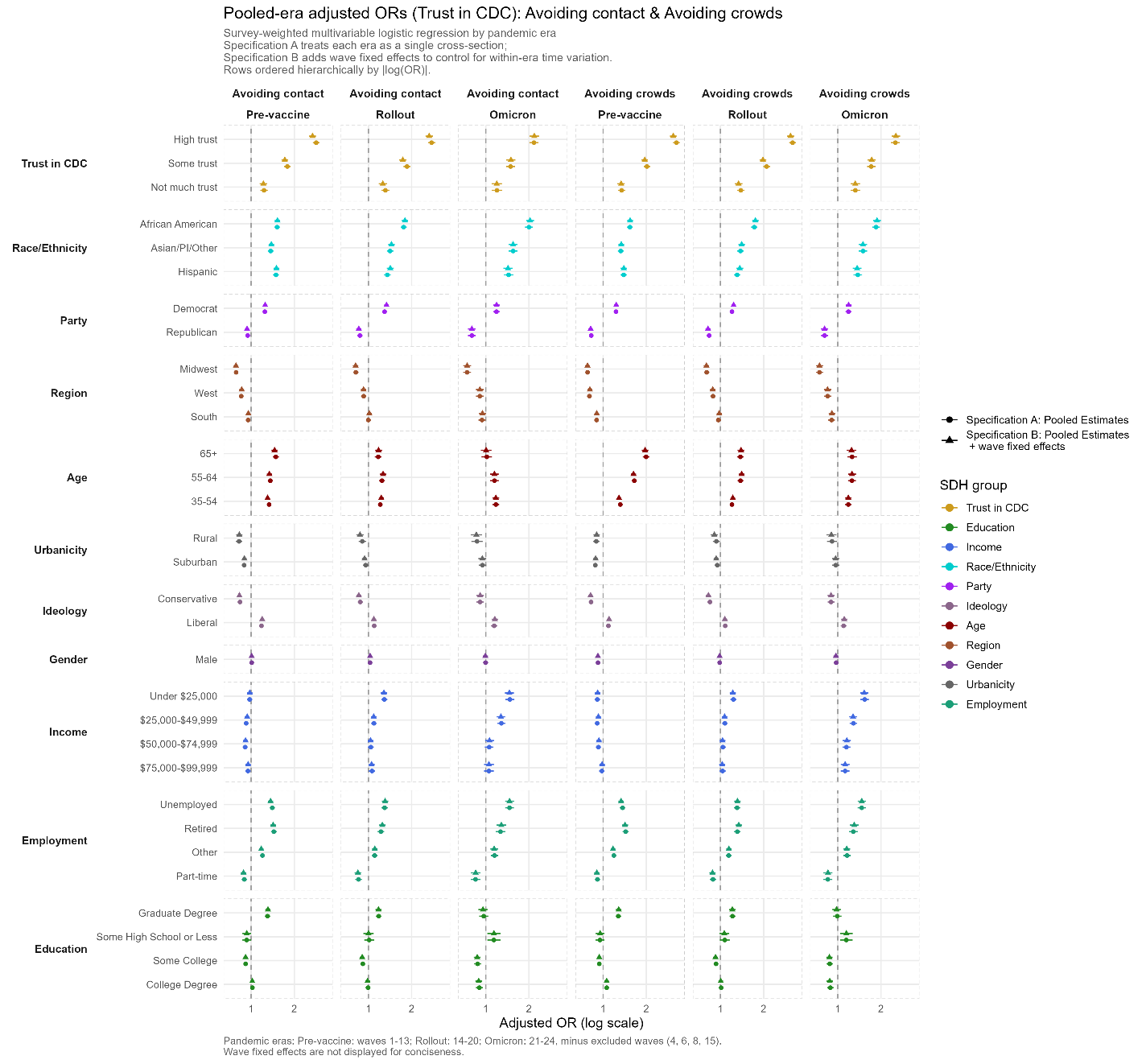

**Fig S12 | Pooled pandemic-era adjusted odds ratios for mask wearing and handwashing, by trust in the CDC and sociodemographic covariates.**

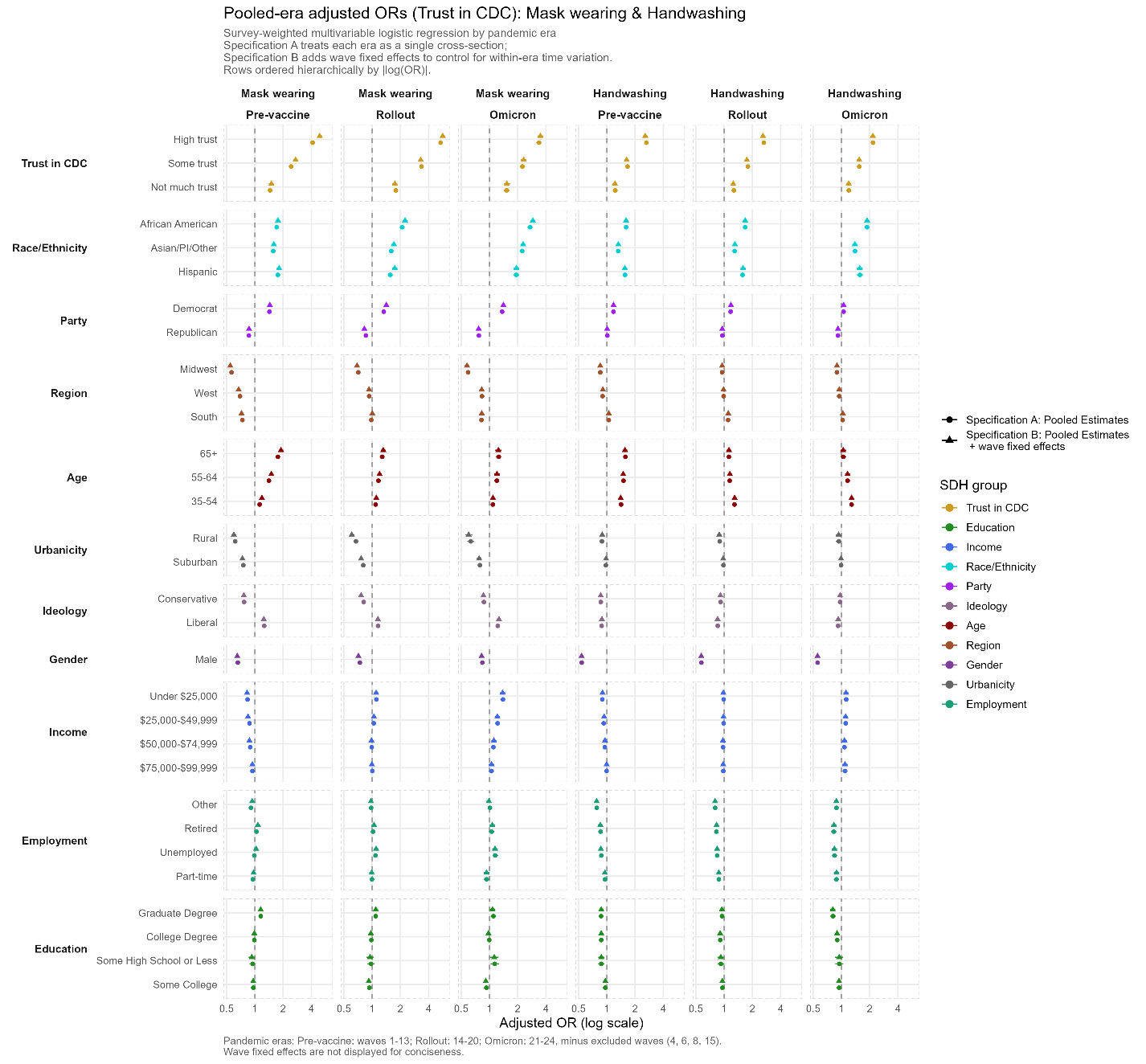

**Fig S13 | Pooled pandemic-era adjusted odds ratios for vaccination (1+ dose(s)) and visiting a doctor or hospital, by trust in the CDC and sociodemographic covariates.**

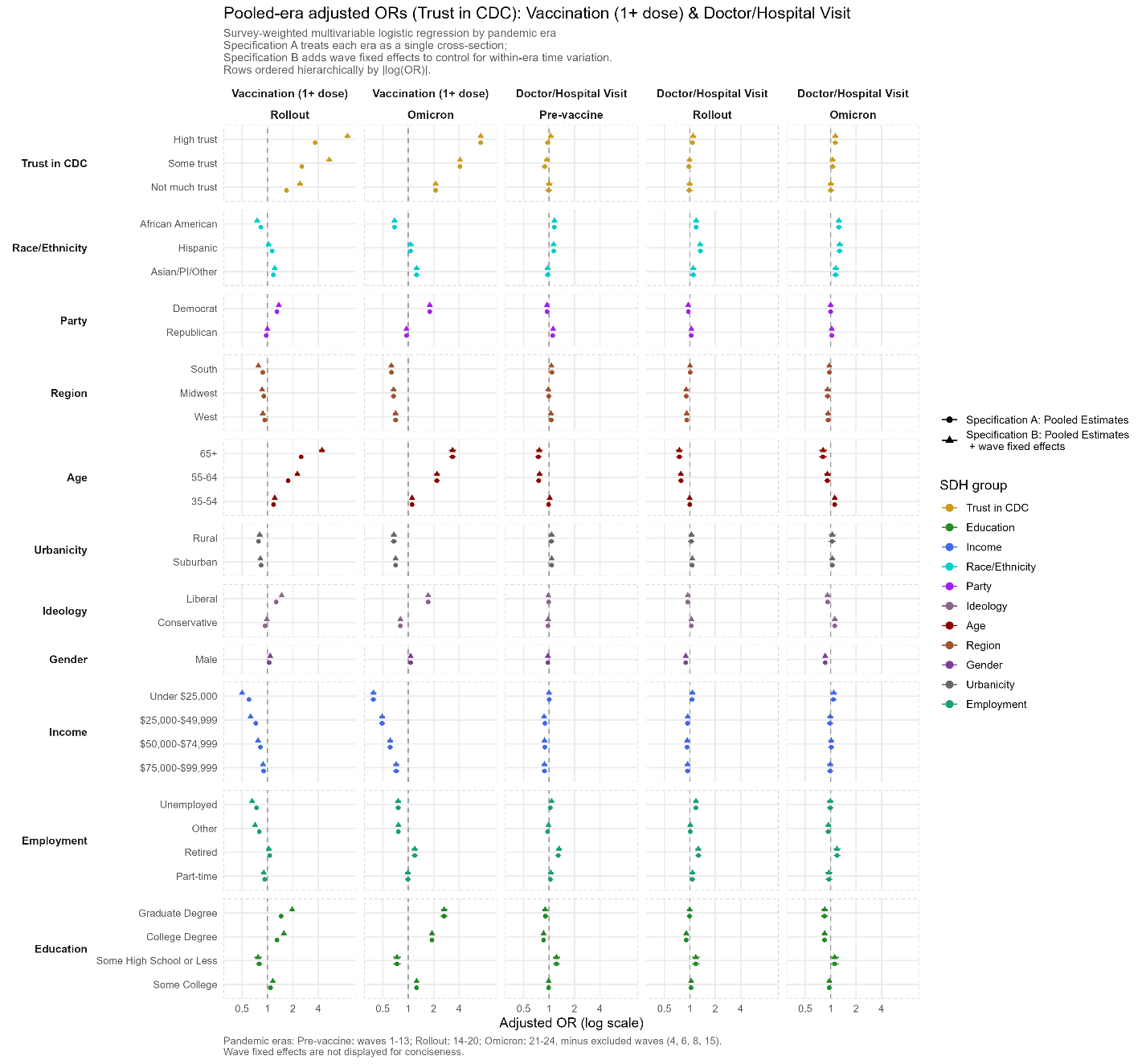

**Fig S14 | Pooled pandemic-era adjusted odds ratios for avoiding contact and avoiding crowds, by trust in banks and sociodemographic covariates.**

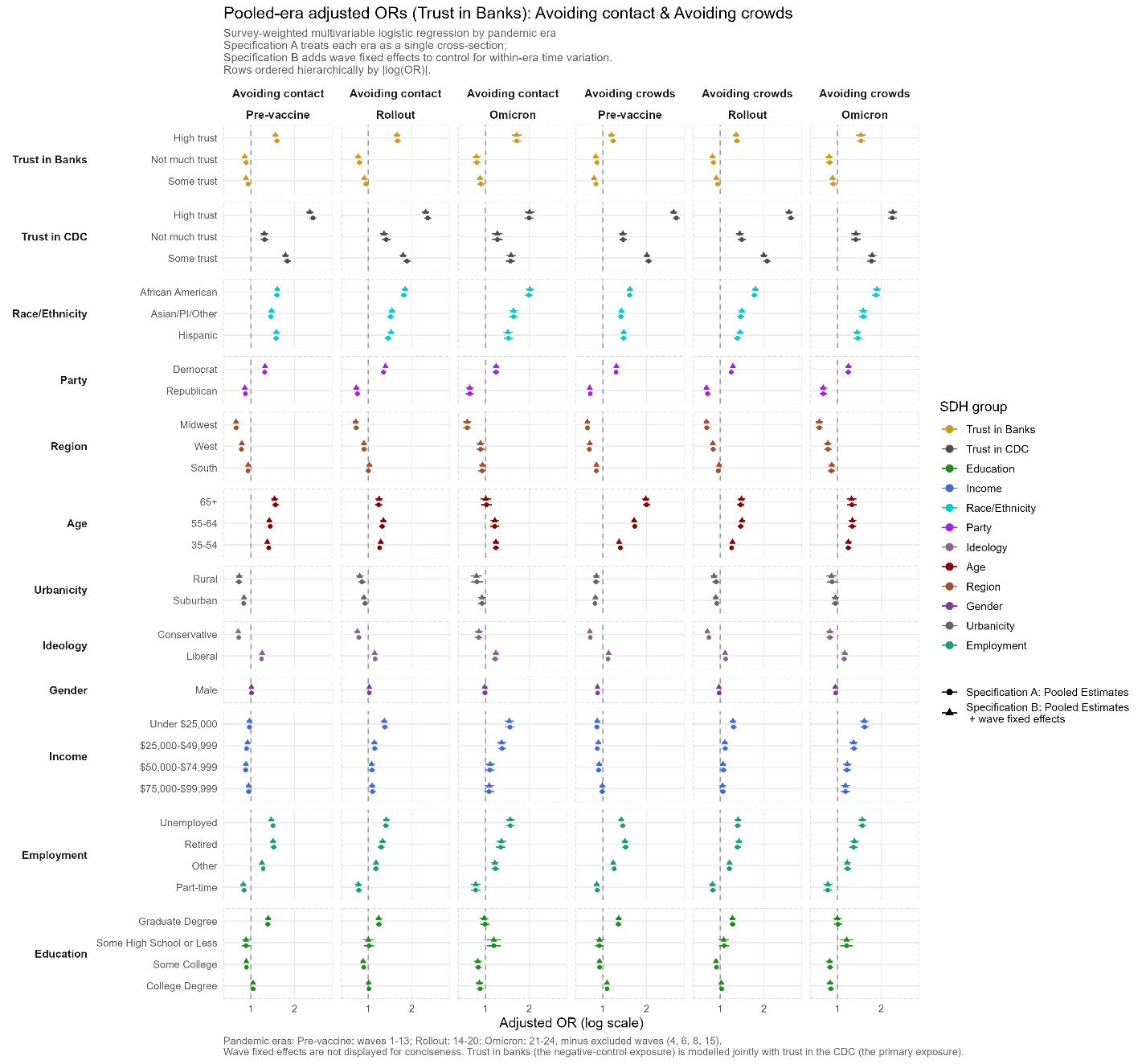

**Fig S15 | Pooled pandemic-era adjusted odds ratios for mask wearing and handwashing, by trust in banks and sociodemographic covariates.**

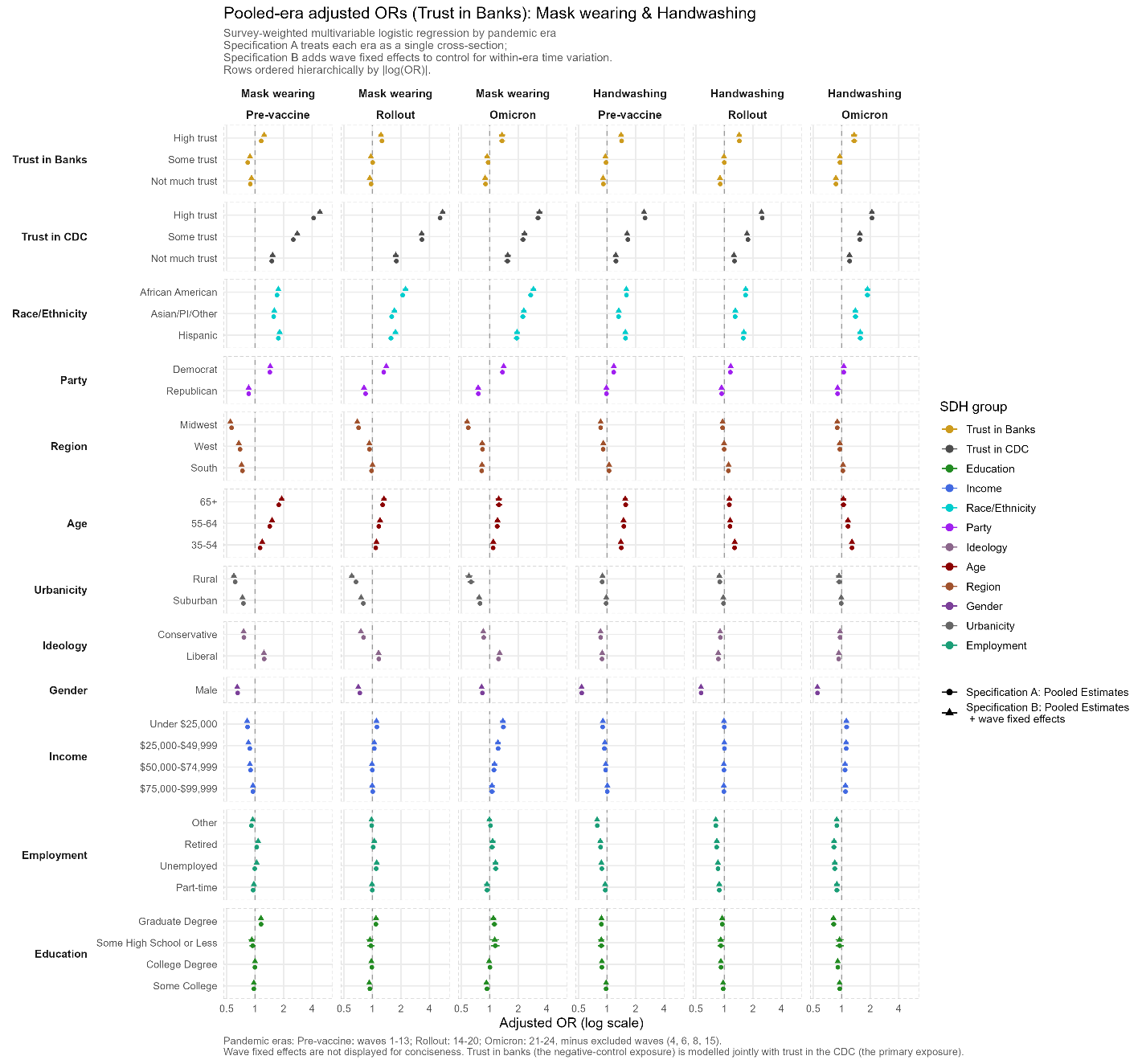

**Fig S16 | Pooled pandemic-era adjusted odds ratios for vaccination (1+ dose(s)) and visiting a doctor or hospital, by trust in banks and sociodemographic covariates.**

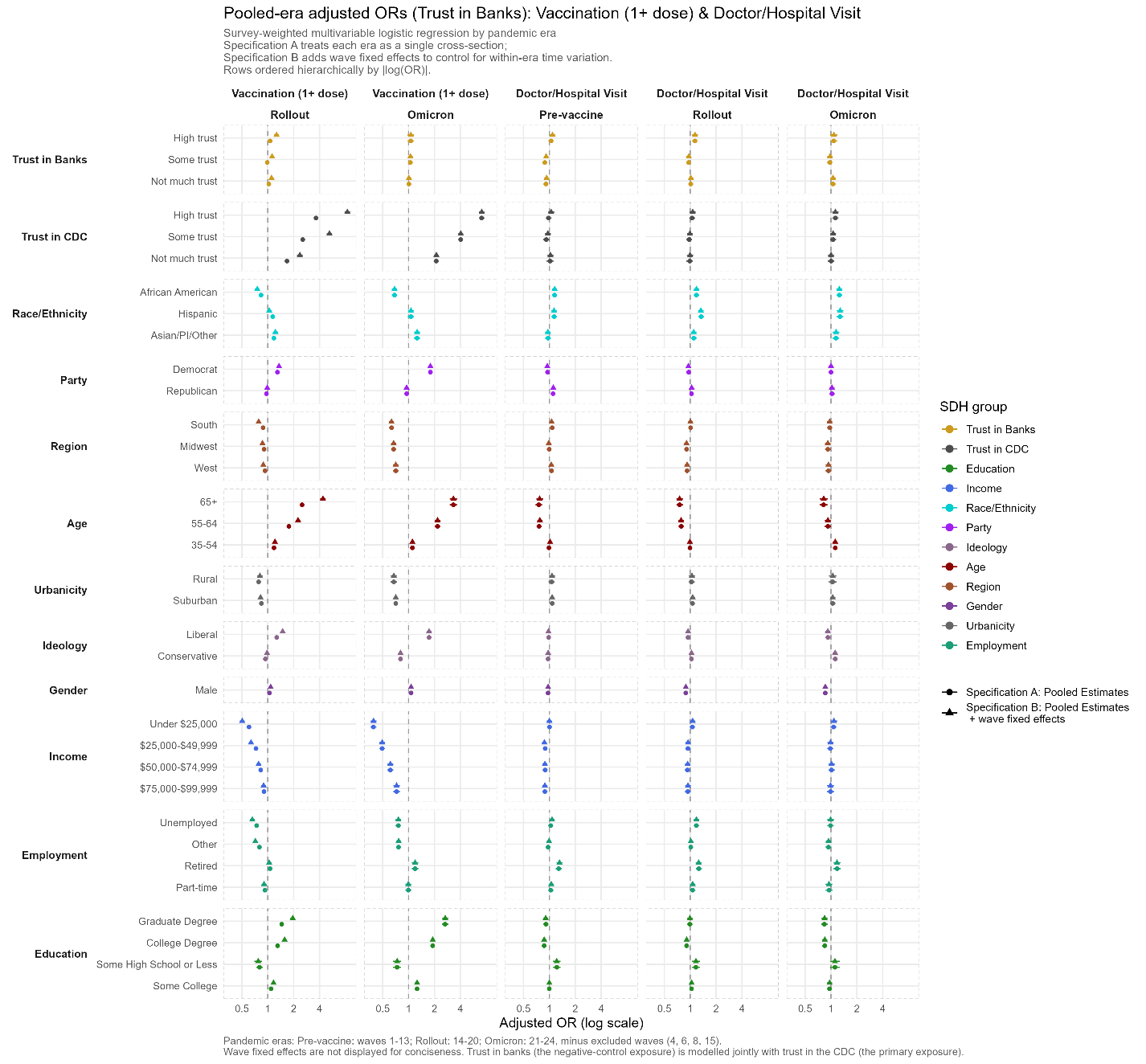

**Fig S17 | Adjusted high- versus no-trust risk differences for trust in the CDC across alternative model specifications**

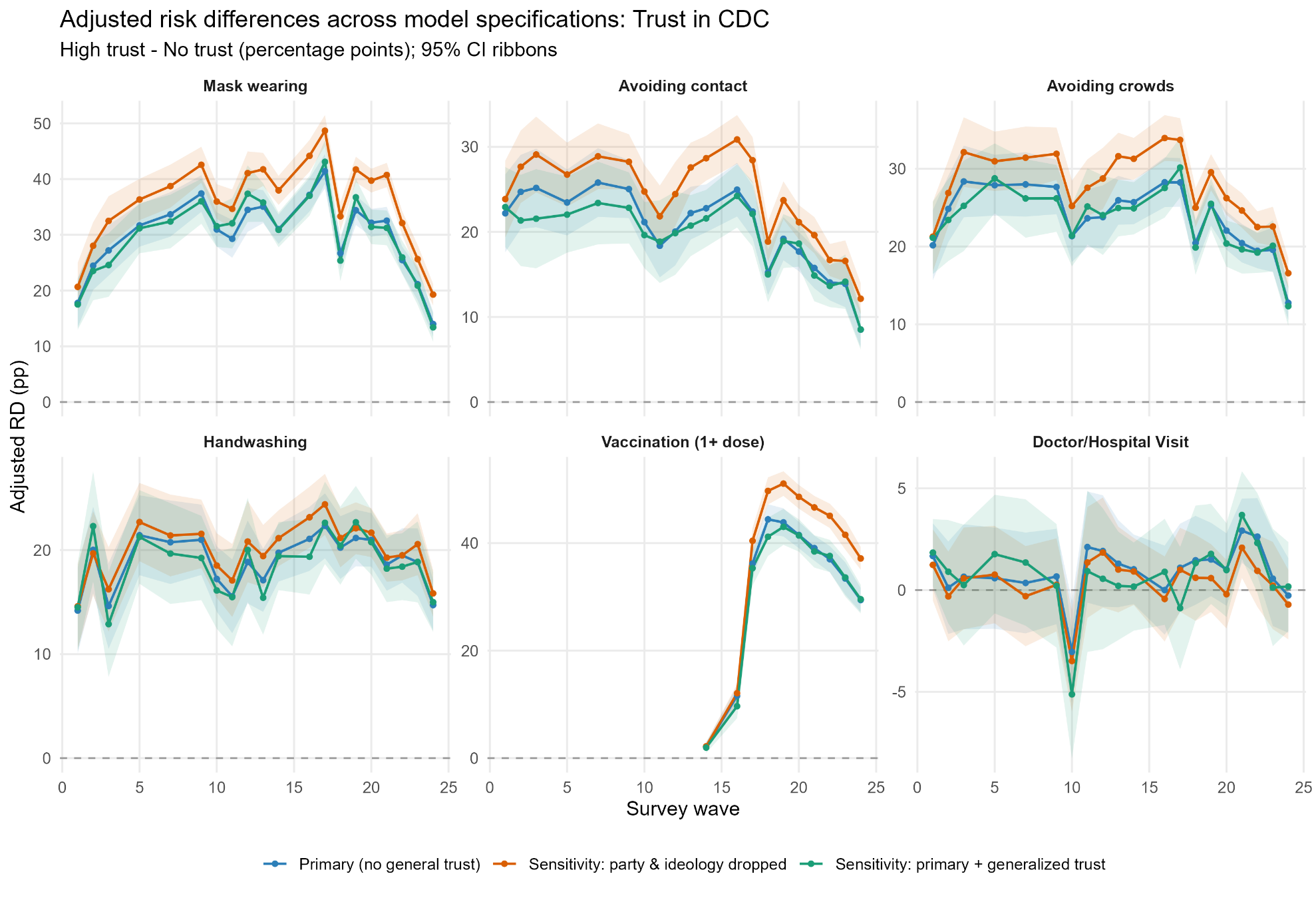

**Fig S18 | Concordance of adjusted trust-in-CDC risk differences with and without adjustment for general trust**

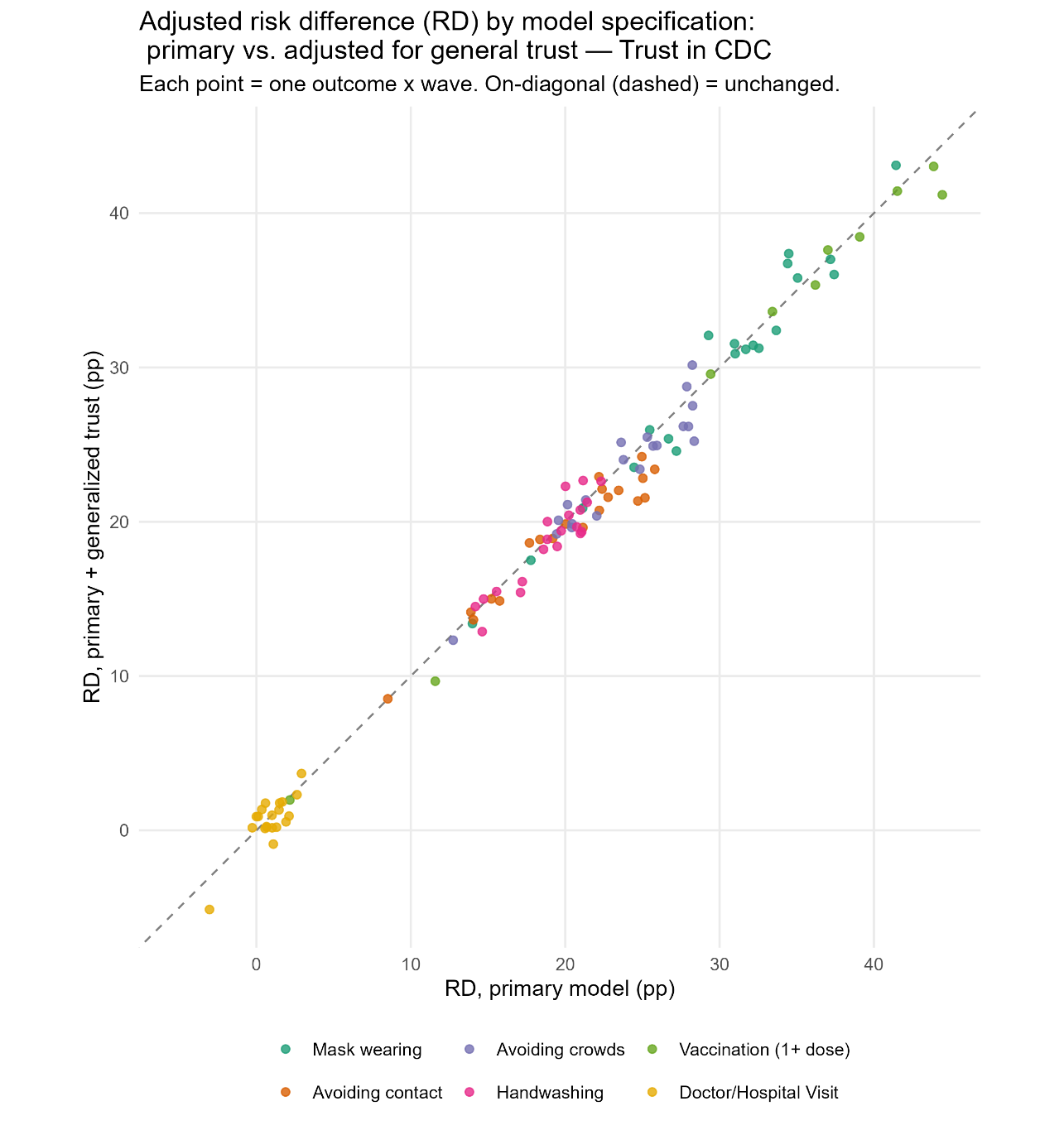

**Fig S19 | Temporal trajectories of institutional trust during the COVID-19 pandemic**

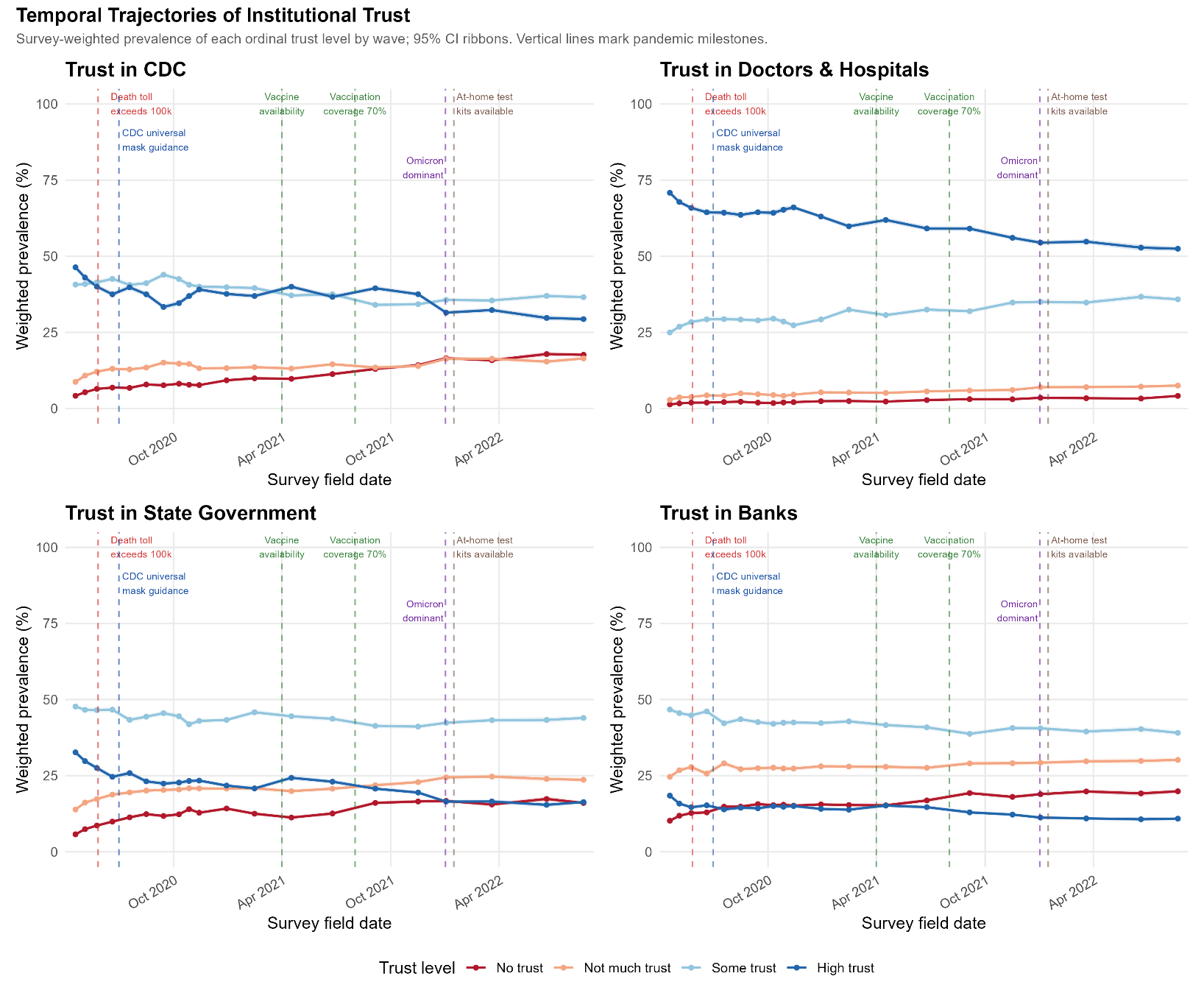
